# Copy number variants differ in frequency across genetic ancestry groups

**DOI:** 10.1101/2024.03.01.24303598

**Authors:** Laura M. Schultz, Alexys Knighton, Guillaume Huguet, Zohra Saci, Martineau Jean-Louis, Josephine Mollon, Emma E.M. Knowles, David C. Glahn, Sébastien Jacquemont, Laura Almasy

## Abstract

Copy number variants (CNVs), which are duplicated or deleted genomic segments larger than 1000 base pairs^1^, have been implicated in a variety of neuropsychiatric and cognitive phenotypes^2-4^. In the first large-scale of examination of genome-wide CNV frequencies across ancestry groups, we found that deleterious CNVs are less prevalent in non-European ancestry groups than they are in European ancestry groups of both the UK Biobank (UKBB) and a US replication cohort (SPARK). We also identified specific recurrent CNVs that consistently differ in frequency across ancestry groups in both the UKBB and SPARK. These ancestry-related differences in CNV prevalence present in both an unselected community population and a family cohort enriched with individuals diagnosed with autism spectrum disorder (ASD) strongly suggest that genetic ancestry should be considered when probing associations between CNVs and health outcomes.

---

CNVs are associated with a wide range of human complex traits^5-7^ and diseases^8-11^. They are especially well-studied in neurodevelopmental^12-15^ and psychiatric disorders^4,15-18^, most notably ASD^19-22^ and schizophrenia^23-25^. Unfortunately, most CNV association studies have been limited to European (EUR) ancestry groups^2,5-9 11^ or pooled across ancestry groups ^10,13-15,17,19-21,23^. While some studies have characterized CNVs in African (AFR)^26-29^ and other non-EUR ancestry groups^15,30-32^, efforts to compare CNV frequencies across ancestry groups have been inconclusive due to limited sample sizes.^14,33-40^ Since genetic variation among human populations^41,42^ contributes to differential disease risk^43-46^, immune response^47,48^, and pharmacogenomics^49,50^, it is plausible that ancestry-related differences in CNV frequency could impact precision medicine. Hence, we took advantage of previously overlooked diversity within the UKBB to compare CNV frequency across four genetic ancestry groups: individuals with inferred EUR (*n* = 51,334), South Asian (SAS; *n* = 8,848), or AFR (*n* = 8,447) ancestry plus a subset of EUR-ancestry individuals who self-identified as “white British” (WB; *n* = 385,636) **(Extended Data Fig. 1)**. Even though the SAS and AFR groups each represent about 2% of the UKBB, they are orders of magnitude larger than the non-EUR groups included in previous CNV studies.

When we considered all autosomal CNVs consisting of at least 50,000 base pairs, we found that the carrier frequency was generally similar across ancestry groups. Nonetheless, there were more deletion (DEL) carriers in the EUR and SAS groups and duplication (DUP) carriers in the WB group than expected given the group size (**Fig. 1a**). For all ancestry groups, DEL carriers were more common than DUP carriers.

**Fig. 1:**
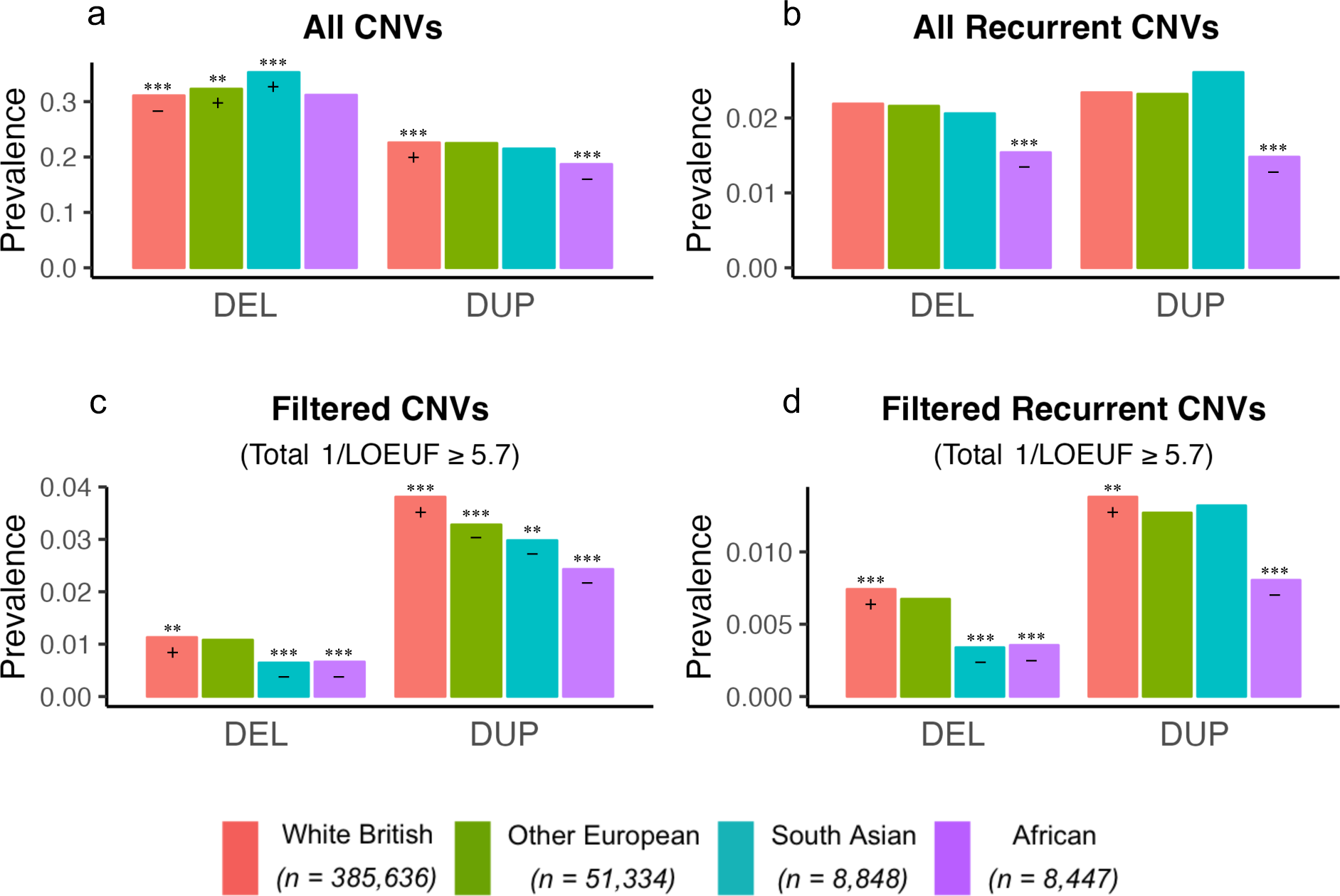
CNV carrier prevalence by ancestry group in the UK Biobank. **a.** When CNVs are not filtered based on recurrence or burden, deletion (DEL) carrier prevalence is higher than expected for the South Asian ancestry group (SAS), and duplication (DUP) carrier prevalence is lower than expected for the African ancestry group (AFR) (*χ*^2^ = 146.7). **b.** When considering only recurrent CNVs, there were fewer AFR DEL and DUP carriers than expected (*χ*^2^ = 48.145). **c.** Filtering instead on burden (total 1/LOEUF ≥ 5.7), there were fewer DEL and DUP carriers than expected for both AFR and SAS (*χ*^2^ = 126.89). **d.** Limiting to carriers of recurrent CNVs with total 1/LOEUF ≥ 5.7, DEL carrier prevalence was lower than expected for both AFR and SAS, but DUP carrier prevalence was only lower than expected for AFR (*χ*^2^ = 61.397). Plus signs indicate significantly higher than expected carrier prevalence, and minus signs indicate significantly lower than expected carrier prevalence. Ancestry-specific carrier prevalence was computed as the number of carriers of at least one DEL (or DUP) divided by the total number of individuals in that ancestry group. Simulated *p*-values (2000 replicates) were less than .0005 for all four chi-square tests of independence. 1/LOEUF, inverse loss-of-function observed/expected upper-bound fraction; **FDR-corrected *p*-value < .005; ***FDR-corrected *p*-value < .0005. An additional category (“Neither”) that was included in each chi-square test is not shown in the bar charts.

Given that CNVs have a range of effect sizes, as judged by their association with health outcomes or by their degree of evolutionary constraint, we considered several subsets of CNV carriers. First, we limited our analyses to carriers of a pre-selected set of recurrent CNVs with common breakpoints that were previously observed in multiple individuals and found to be associated with neuropsychiatric phenotypes (**Supplementary Table 1**). We observed 50 unique recurrent deletions (**Supplementary Table 2**) and 60 unique recurrent duplications (**Supplementary Table 3**) at these loci in the UKBB. Of the 110 unique recurrent CNVs observed in the combined sample, 106 were present in the WB. The 4 CNVs not observed in the WB group were present in the EUR group, where we observed 77 unique recurrent CNVs. The 40 unique recurrent CNVs observed in the AFR group and the 36 in the SAS group were subsets of those observed in the WB and EUR groups; none of the 110 recurrent CNVs were unique to the non-EUR ancestry groups in the UKBB. There were significantly fewer AFR-ancestry recurrent DEL and DUP carriers than expected under the null hypothesis that recurrent CNV carrier frequency is independent of ancestry group (**Fig. 1b**).

Next, we filtered CNV carriers using the loss-of-function observed/expected upper bound fraction (LOEUF)^51^ constraint metric. When we limited our analyses to carriers of CNVs with constraint scores equivalent to disruption of at least two predicted loss-intolerant genes (i.e., total 1/LOEUF ≥ 5.7 summed across the genes within the CNVs carried by an individual), ancestry-related differences persist and become even stronger. Consistent with previous evidence that DELs in coding sequences are under stronger purifying selection than DUPs^52^, the burden-filtered carrier frequency for DELs was substantially lower than that for DUPs for all four ancestry groups. The deleterious DEL carrier frequency for WB individuals was nearly twice that of AFR or SAS individuals, both when we filtered by 1/LOEUF summed across all CNVs (**Fig. 1c**) and when we limited the summation to recurrent CNVs (**Fig. 1d**). Likewise, carriers of deleterious DUPs were more prevalent in the WB group and less prevalent in the AFR group than expected. We obtained similar results when we excluded all individuals related at the third-degree or closer (**Extended Data Fig. 2; Supplementary Table 4**).

We also examined the prevalence of individual recurrent CNVs. To maximize power, we limited our analysis to the 11 recurrent CNVs (5 DELs and 6 DUPs) that each had a total of 275 or more observations across the four ancestry groups. We found ancestry-related differences in the prevalence of 6 of these CNVs (**Fig. 2**), with some CNVs being more prevalent in WB individuals but others occurring at higher rates in the AFR and SAS groups. While there were some related individuals among the carriers of some of the CNVs, the pattern of frequency differences was essentially unchanged when we excluded all individuals with third-degree or closer relatives from the dataset (**Extended Data Fig. 3; Supplementary Table 4**). Hence, the observed differences cannot be explained by related individuals carrying the same CNVs.

**Fig. 2:**
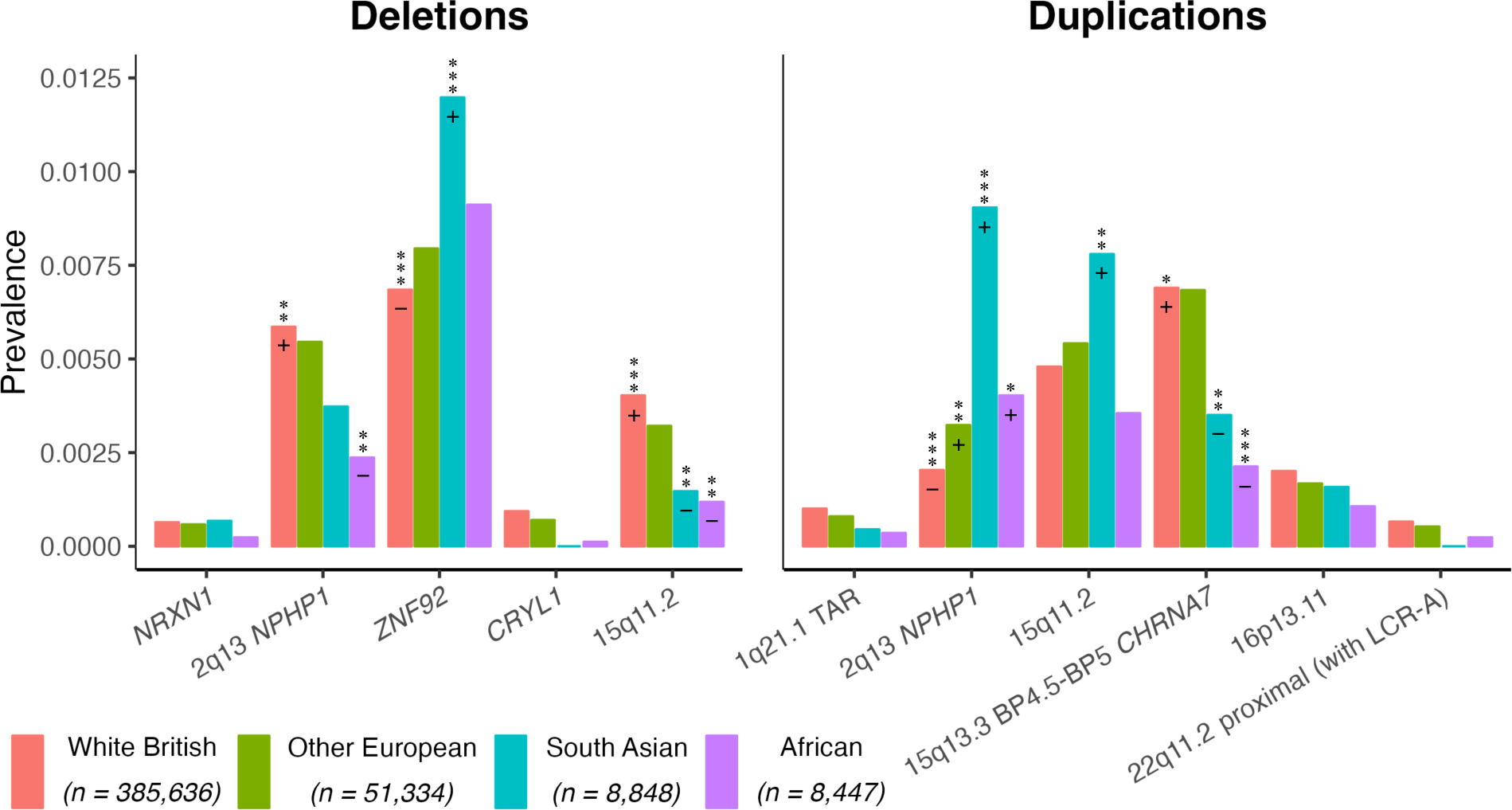
Prevalence of individual recurrent CNVs across ancestry groups in the UK Biobank. There were significant differences between expected and observed counts for 6 out of the 11 recurrent CNVs selected for analysis, *χ*^2^ = 425.3, simulated *p*-value < .0005 (2000 replicates). Carrier prevalence was calculated as the number of carriers of a given recurrent CNV divided by the total number of individuals in that ancestry group. Plus and minus signs indicate that the standardized residuals were statistically significantly higher or lower than zero, respectively, after FDR correction. *FDR-corrected *p*-value < .05; **FDR-corrected *p*-value < .005; ***FDR-corrected *p*-value < .0005. An additional category (“none of the above”) that was included in the chi-square test is not shown in the bar chart.

Ascertainment bias is another potential explanation for the lower rate of deleterious CNVs in some ancestry groups within the UKBB. Immigrants may be less likely to participate in the UKBB, especially if they are carriers of deleterious CNVs. While EUR, AFR, and SAS individuals were more likely to have been born outside the UK or Ireland than WB individuals, the proportions of SAS and AFR individuals who were immigrants did not significantly differ for recurrent CNV carriers compared to non-carriers (**Extended Data Fig. 4**), suggesting that differential immigration rates cannot explain the differences in CNV prevalence across ancestry groups.

Demographic differences between the UKBB ancestry groups could also contribute to the observed differences in CNV prevalence. Individuals in the SAS and AFR ancestry groups tend to be younger than those in the WB and EUR groups, and the female-to-male ratios differed across the ancestry groups (**Supplementary Tables 5-6**). Furthermore, median Townsend deprivation index scores suggest that there are socioeconomic differences that could be associated with ancestry, sex, and recurrent CNV carrier status (**Supplementary Table 7**). We used propensity score matching to balance the sample sizes and control for the potentially confounding effects of these variables on CNV carrier rates by down-sampling the WB group to create two subgroups matched to the AFR and SAS groups (**Extended Data Fig. 5; Supplementary Tables 8-9**). The AFR-ancestry group had significantly lower odds of carrying both unfiltered and 1/LOEUF-filtered recurrent DELs and DUPs than its matched WB group (all two-sided Fisher’s exact test *p*-values < .0000005; **Fig. 3**). Indeed, the matched comparisons yielded more pronounced differences than the unmatched ones despite having smaller sample sizes. In contrast, differences between the SAS and matched WB group were somewhat attenuated. Nonetheless, the SAS group showed significantly lower odds of carrying 1/LOEUF-filtered recurrent DELs when compared to its matched WB group (*p* < .00001).

**Fig. 3:**
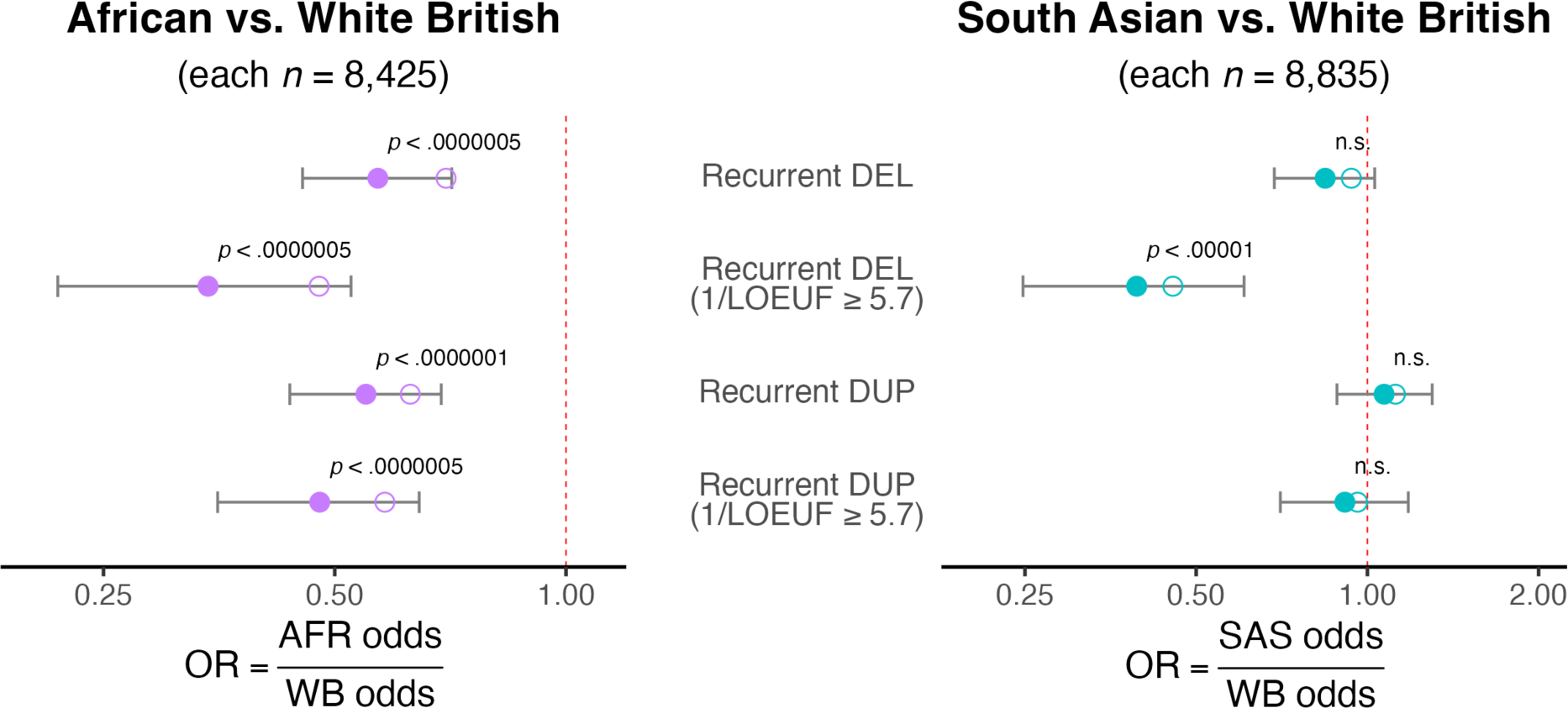
Odds of carrying deleterious CNVs in AFR- and SAS-ancestry individuals compared to WB individuals in the UK Biobank after propensity-score matching on Townsend deprivation index, age, and sex. Odds were computed as the number of carriers divided by the number of non-carriers of a given type of recurrent CNV. Odds ratios (ORs) were computed as AFR odds divided by WB odds (purple dots) or SAS odds divided by WB odds (blue dots) using unmatched (open dots) or matched (filled dots) data. Error bars indicate 95% Fisher confidence limits for the OR computed using matched data, and the *p*-value is for the corresponding two-sided Fisher’s exact test for that OR. The red dashed lines indicate the expected OR of 1 (i.e., equal odds). AFR odds were lower than the WB odds at each level of filtering, but SAS odds were not consistently lower than the WB odds. Unmatched odds were calculated using data from 385,636 White British (WB), 8,447 African (AFR), and 8,848 South Asian (SAS) individuals.

We also used the matched datasets to compare the odds of carrying our 11 recurrent CNVs of interest (**Fig. 4**) and found that most of the observed ancestry-related differences in CNV prevalence remained significant. Using two-sided Fisher’s exact tests of the carrier ORs, we found that the AFR group had significantly lower odds of carrying 2q13 *NPHP1* DEL, *CRYL1* DEL, 15q11.2 DEL, 15q13.3 BP4.5-BP5 *CHRNA7* DUP, 16p13.11 DUP, and 22q11.2 proximal (with LCR-A) DUP as well as higher odds of carrying 2q13 *NPHP1* DUP compared to its matched WB group. The SAS group also had significantly lower odds of carrying 2q13 *NPHP1* DEL, *CRYL1* DEL, 15q11.2 DEL, 15q13.3 BP4.5-BP5 *CHRNA7* DUP, and 22q11.2 proximal (with LCR-A) DUP and higher odds of carrying 2q13 *NPHP1* DUP compared to its matched WB group. Additionally, the SAS group had higher odds of carrying *ZNF92* DEL and 15q11.2 DUP compared to its matched WB group.

**Fig. 4:**
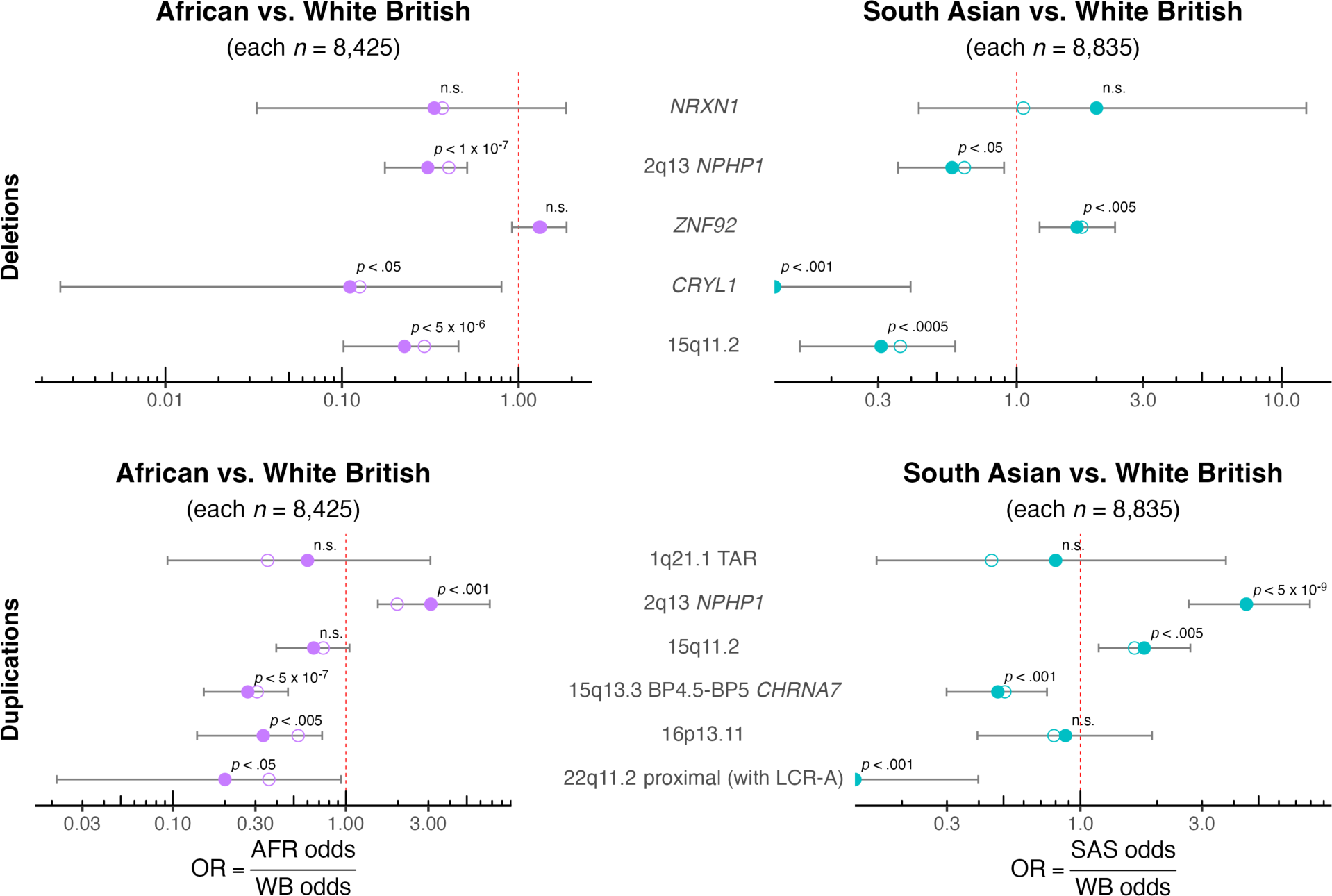
The odds of carrying individual recurrent CNVs for non-EUR compared to WB ancestry groups in the UK Biobank. Odds were computed as the number of carriers divided by the number of non-carriers of a given recurrent CNV. Odds ratios (ORs) were computed as AFR odds divided by WB odds (purple dots) or SAS odds divided by WB odds (blue dots) using unmatched (open dots) or matched (filled dots) data. Error bars indicate 95% Fisher confidence limits for the OR computed using matched data, and the *p*-value is for the corresponding two-sided Fisher’s exact test for that OR. The red dashed lines indicate the expected OR of 1 (i.e., equal odds). Compared to the WB odds, AFR odds were significantly lower for 6 and significantly higher for 1 of the 11 selected recurrent CNVs. SAS odds were significantly lower than WB odds for 5 of the same 6 recurrent CNVs and significantly higher than WB for 3 recurrent CNVs, including the same one observed to be higher for AFR. The two blue half circles correspond to ORs of zero; neither of those recurrent CNVs were observed in the SAS ancestry group. Unmatched odds were calculated using data from 385,636 White British (WB), 8,447 African (AFR), and 8,848 South Asian (SAS) individuals.

Finally, we replicated our study using SPARK, a younger United States cohort enriched with individuals diagnosed with ASD and ID (**Supplementary Tables 10-12**). We observed 51 unique recurrent deletions (**Supplementary Table 13**) and 51 unique recurrent duplications (**Supplementary Table 14**) across the three largest inferred ancestry groups (**Extended Data Fig. 6**) comprised of 46,869 EUR, 7,870 admixed American (AMR), and 3,680 AFR individuals. We used propensity score matching to balance the sample sizes and control for potentially confounding effects of ASD and ID status, age, and sex on recurrent CNV carrier rates by down-sampling the EUR group to create two subgroups matched to the AFR and AMR groups (**Extended Data Fig. 7; Supplementary Tables 15-16**). Replicating our UKBB results, we found that the SPARK AFR group had significantly lower odds of carrying both unfiltered and 1/LOEUF-filtered recurrent DELs and DUPs than its matched EUR group (all two-sided Fisher’s exact test *p*-values < .005; **Fig. 5**). The AMR group had significantly lower odds of carrying both unfiltered and 1/LOEUF filtered recurrent DUPs (two-sided Fisher’s exact test *p*-values < .005; **Fig. 5**).

**Fig. 5:**
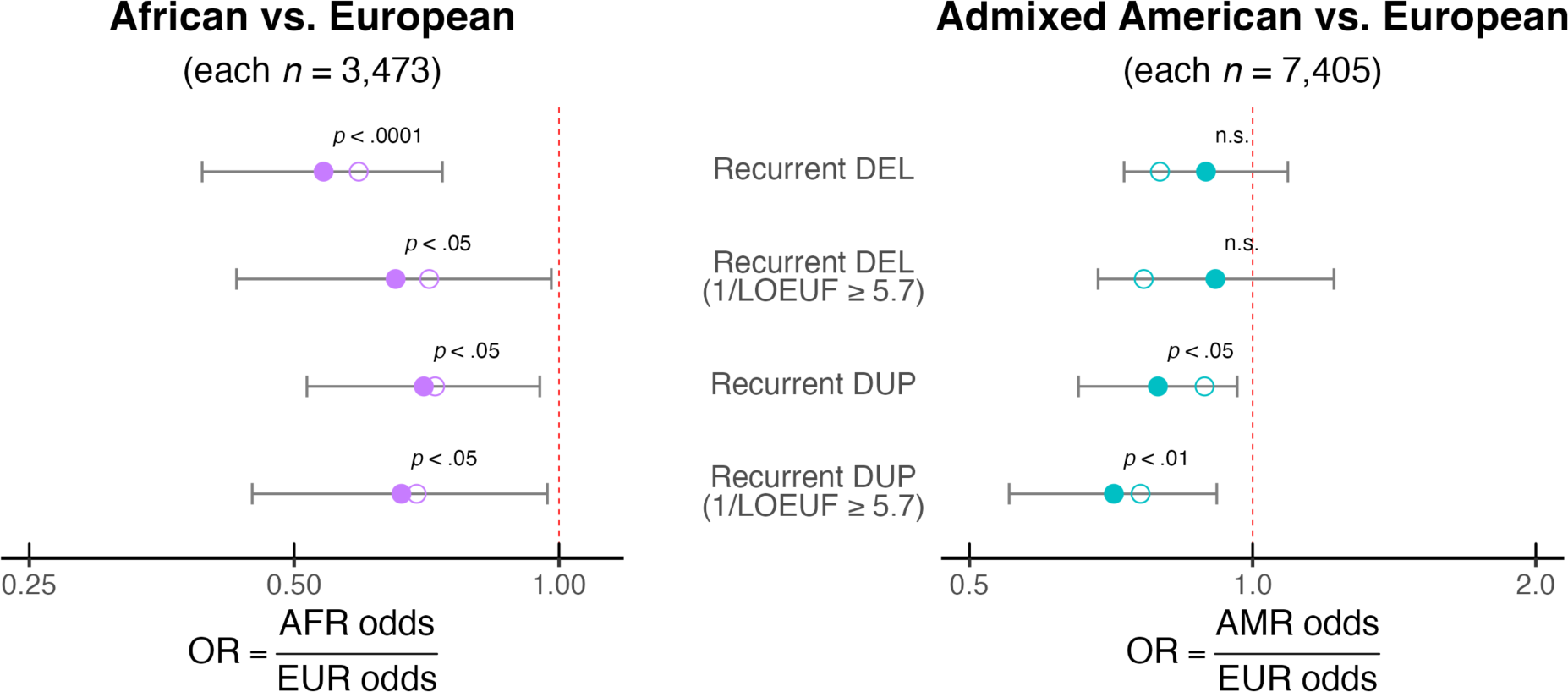
Odds of carrying deleterious CNVs in AFR- and AMR-ancestry individuals compared to EUR-individuals in SPARK after propensity-score matching on autism spectrum disorder and intellectual disability status, age, and sex. Odds were computed as the number of carriers divided by the number of non-carriers of a given type of recurrent CNV. Odds ratios (ORs) were computed as AFR odds divided by EUR odds (purple dots) or AMR odds divided by WB odds (blue dots) using unmatched (open dots) or matched (filled dots) data. Error bars indicate 95% Fisher confidence limits for the OR computed using matched data, and the *p*-value is for the corresponding two-sided Fisher’s exact test for that OR. The red dashed lines indicate the expected OR of 1 (i.e., equal odds). AFR carrier odds were lower than the EUR odds at each level of filtering, but AMR odds were significantly lower only for recurrent DUP carriers. Unmatched odds were calculated using data from 46,869 European (EUR), 3,680 African (AFR), and 7,870 admixed American (AMR) individuals.

We also used the matched datasets to compare the odds of carrying the same 11 recurrent CNVs we analyzed for the UKBB along with two additional recurrent CNVs that had at least 75 copies each in SPARK despite not meeting our inclusion criteria for the UKBB (**Fig. 6**). Replicating our UKBB results, the SPARK AFR group showed significant differences for 2q13 *NPHP1* DEL, 15q11 DEL, 2q13 *NPHP1* DUP, 15q13.3 BP4.5-BP5 *CHRNA7* DUP, and 16p13.11 DUP carrier odds relative to its matched EUR group (two-sided Fisher’s exact test *p*-values < .05). Additionally, the SPARK AFR group had significantly lower odds of carrying *NRXN1* DEL and higher odds of carrying 16p12.1 DEL relative to its matched EUR group, and the SPARK AMR group had significantly lower odds of carrying *NRXN1* DEL, 15q11.2 DEL, and 1q21 TAR DUP relative to its matched EUR group (two-sided Fisher’s exact test *p*-values < .05).

**Fig. 6:**
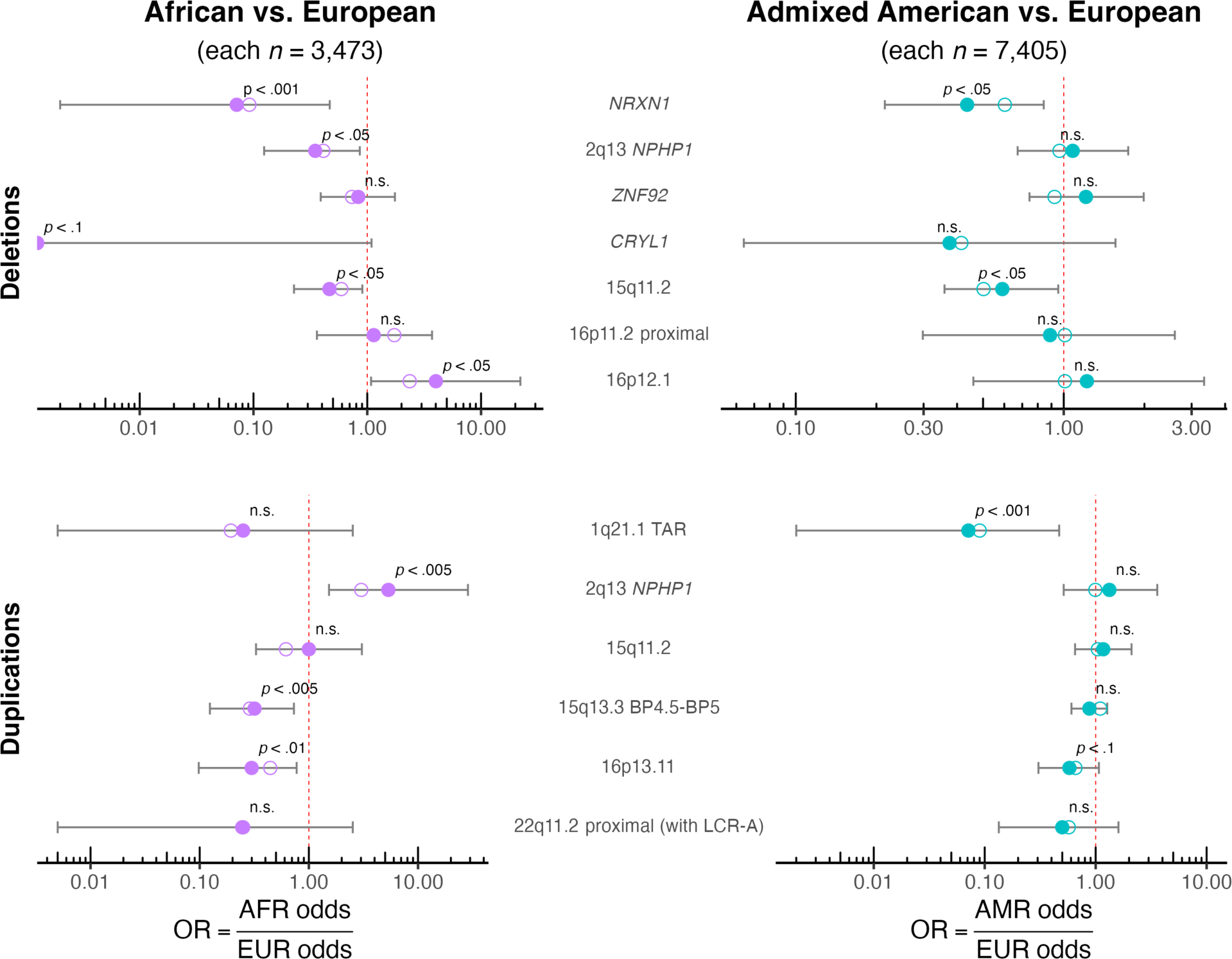
Odds of carrying individual recurrent CNVs for non-EUR compared to EUR ancestry groups in SPARK. Odds were computed as the number of carriers divided by the number of non-carriers of a given recurrent CNV. Odds ratios (ORs) were computed as AFR odds divided by EUR odds (purple dots) or AMR odds divided by EUR odds (blue dots) using unmatched (open dots) or matched (filled dots) data. Error bars indicate 95% Fisher confidence limits for the OR computed using matched data, and the *p*-value is for the corresponding two-sided Fisher’s exact test for that OR. The red dashed lines indicate the expected OR of 1 (i.e., equal odds). The purple half circle corresponds to an OR of zero; this recurrent CNV was not observed in the AFR ancestry group. Unmatched odds were calculated using data from 46,869 European (EUR), 3,680 African (AFR), and 7,870 admixed American (AMR) individuals.

One potential explanation for the difference in prevalence of specific CNVs across ancestry groups is that population variation in the flanking sequence may affect the probability of deletions and duplications in a region. For example, chromosomal regions with polymorphic inversions have been shown to be enriched for recurrent CNVs associated with developmental delay and neuropsychiatric disorders.^53^

We demonstrated that ancestry-related differences in CNV carrier prevalence are present in both unselected community populations (UKBB) and cohorts enriched with ASD-diagnosed individuals (SPARK). We replicated the observed differences between the AFR and WB cohorts in the UKBB by comparing the AFR and EUR cohorts in SPARK, which is notable given the ascertainment differences, differing genotyping platforms (**Methods**), and presumed genetic differences between the homogeneous WB subset of the UKBB and a EUR-ancestry cohort from the United States^54^. Furthermore, SAS (UKBB) and AMR (SPARK) ancestry groups also exhibited unique patterns of CNV prevalence, demonstrating that differences in CNV carrier prevalence cannot be generalized as “EUR vs. non-EUR” differences.

Given that African populations have been shown to have greater genetic diversity^55^, the finding of fewer rare CNVs in the AFR groups of UKBB and SPARK is somewhat surprising. One possible explanation for our ancestry-divergent results could be the “Euro-centric” focus of previous studies of genetic variants.^43^ To focus on deleterious CNVs, we limited some of our analyses to recurrent CNVs that had been previously implicated in neuropsychiatric disorders and/or filtered our results based on the LOEUF metric. Our targeted recurrent CNVs were originally identified in cohorts dominated by EUR-ancestry individuals, so it is conceivable that there are as-yet-undiscovered CNVs with medical relevance that are more common in other ancestry groups. Also, the LOEUF constraint metric was derived from the gnomAD v2 reference database, which includes sequences from 64,603 individuals from European populations and only 12,487 individuals from African (or African-American) and 15,308 individuals from South Asian populations^51^. We expect that the discovery of additional recurrent CNVs, especially those smaller than 50 kb, will be facilitated by the increasing availability of long-read whole genome sequence data collected from diverse populations, such as *All of Us*^40,56^. In any case, it is likely that differing linkage disequilibrium patterns^35^ and flanking sequences^57^ also contributed to the ancestry-related differences in CNV frequency that we found. Future CNV studies utilizing long-read sequences should clarify the extent to which polymorphic differences in the flanking sequence across populations contribute to the observed differences in CNV frequency.

These findings are limited by the sample sizes of the AFR, SAS, and AMR ancestry groups in the UKBB and SPARK, which are much smaller than those of the WB and EUR ancestry groups. Larger CNV studies of diverse samples are needed. Additional limitations include our focus on only CNVs over 50 kilobases, a by-product of using genotype array data to call CNVs, and the choice of LOEUF as a metric of the deleteriousness of a CNV. Given that LOEUF is focused on coding genes, non-coding CNVs that impact important regulatory elements may have been missed in our filtered set of deleterious CNVs with 1/LOEUF ≥ 5.7. Strengths of the study include the use of propensity score matching to address potential sources of bias and the consistency of results across samples with very different demographic profiles and ascertainment schemes.

Although classifying individuals into continental ancestry groups imposes discrete categories onto what is actually a continuum of human genetic variation^58,59^, it has nonetheless been a useful approach for considering the potential effects of population structure on genome-wide association analyses^60,61^. We recommend that CNV association studies also be adjusted for population structure, ideally in a quantitative way that reflects the continuous spectrum of genetic variation, to limit the risk of spurious discoveries.

## Methods

### Cohorts

We obtained de-identified genotype and phenotype data for the UK Biobank^62^ (UKBB; application number 40980), a prospective population-based cohort of approximately 500,000 United Kingdom residents who were 40-69 years old when they were recruited between 2006 and 2010.

We replicated our primary results using data from Simons Powering Autism Research for Knowledge (SPARK)^63^, a United States cohort of children and dependent adults diagnosed with autism spectrum disorder (ASD) and their families.

### Genotyping and CNV Calling

UKBB DNA samples were extracted from blood and genotyped on either the UK BiLEVE Axiom Array (*n* = 49,950) or the UK Biobank Axiom Array (*n* = 438,427) by Affymetrix. We selected 733,256 probes shared by the two arrays when mapped to hg19. Intensity data were used to call CNVs for the 459,855 individuals who remained after excluding 28,522 samples that failed to meet our quality control (QC) standards of genotype call rate > 0.95, |waviness factor| < 0.05, log *R* ratio SD < 0.35, and B allele frequency SD < 0.08.

SPARK DNA samples were extracted from saliva and genotyped on four different Illumina Infinium arrays: Global Screening Array (GSA)-24 v1.0 (*n* = 26,868), GSA-24 v2.0 (*n* = 32,397), CoreExome-24 v1.1 (*n* = 1,382), and CoreExome-24 v1.3 (*n* = 6,024). When mapped to hg19, the two GSA arrays had 617,394 autosomal SNPs in common with each other and 218,457 SNPs in common with the 733,256 probes used to call CNVs for the UKBB, and the CoreExome arrays had 514,277 autosomal SNPs in common with each other and 98,648 in common with the UKBB probes. The four Illumina arrays had a total of 514,273 autosomal SNPs in common. Intensity data were used to call CNVs for the 65,425 individuals who remained after excluding the samples that failed to meet the same QC standards listed above for the UKBB.

CNVs were called in parallel by PennCNV^64^ and QuantiSNP^65^ using our previously published pipeline^66,67^. Both algorithms combine normalized intensity data with log R ratio (LRR) and B allele frequency (BAF) into hidden Markov models to detect CNVs that meet the following criteria: coverage ≥ 3 consecutive probes, size ≥ 1 kB, and confidence score ≥ 15. CNVs jointly detected by both algorithms were merged using CNVision^68^, and a CNV inheritance algorithm was used to concatenate adjacent duplications (DUPs) or deletions (DELs) separated by a gap no larger than 150 kB. Only CNVs meeting the following criteria were selected for further analyses: confidence score ≥ 30 for at least one detection algorithm; size ≥ 50 kB; unambiguous type (DUP or DEL); and less than 50% overlap with segmental duplicates, HLA regions, or centromeric regions.

### Genetic Ancestry Inference

Ten principal components (PCs) were computed from the imputed genotypes supplied by the UKBB (Data-Field 22828; v.3) and projected onto the 1000 Genomes PC space using KING v2.2.4^69^. Genetic ancestry was inferred from these PCs via a support vector machine algorithm using the e1071 R package^70^. Additionally, a subset of 385,636 European-ancestry individuals who self-identified as being “white British” (Data-Field 22006) was defined; the white British (WB) subgroup formed a tight subcluster within the European-ancestry cluster on the PC2 vs. PC1 plot (**Extended Data Fig. 1a**). All subsequent analyses were limited to these WB individuals in addition to 51,334 other European (EUR), 8,848 South Asian (SAS), and 8,447 African (AFR) ancestry individuals. While KING called the ancestry whenever the posterior probability was at least 65%, the majority (93.2%) of the calls were made with posterior probability ≥ 90% (**Extended Data Fig. 1b**), suggesting that admixture was limited. Nonetheless, we recognize that genetic ancestry is continuous and does not equate with self-identified ethnicity (Data-Field 21000; **Extended Data Fig. 1c**).

Genetic ancestry was inferred using this same approach for the SPARK samples included in the iWESv2 or the WGS1-3 data releases. After excluding any individuals who were missing from the *individuals_registration* file supplied with the July 2023 SPARK V10 release, we limited all subsequent analyses to 46,869 EUR, 7,870 admixed American (AMR) and 3,680 AFR ancestry individuals (**Extended Data Fig. 6; Supplementary Tables 10-12**).

### CNV Annotation

CNVs were annotated in GRCh37 using GENCODE Release 35 (https://www.gencodegenes.org/human/release_35lift37.html) and Ensembl (https://grch37.ensembl.org/index.html). Genes that were fully contained within a CNV were identified, and the inverse loss-of-function observed/expected upper-bound fraction (1/LOEUF) from gnomAD (version 2.1.1)^51^ was summed across all full genes contained within all CNVs detected for each study participant. A CNV was classified as a “recurrent CNV” if it met defining criteria for any of a pre-selected set of recurrent and single-gene autosomal CNVs that have been previously associated with neurodevelopmental or neuropsychiatric phenotypes (**Supplementary Table 1**). Partial genes were also included in the 1/LOEUF calculations for these recurrent CNVs. We use “all CNVs” to refer to these pre-selected recurrent CNVs plus all other CNVs over 50kB. An individual’s total 1/LOEUF was summed across full genes for all CNVs as well as any partial genes that were included in the pre-defined recurrent CNVs, whereas total 1/LOEUF calculations made for an individual’s recurrent CNVs were limited to the full and partial genes present within these pre-defined recurrent and single-gene CNVs. We observed 50 unique recurrent DELs (**Supplementary Table 2**) and 60 unique recurrent DUPs (**Supplementary Table 3**) across our four defined UKBB ancestry groups. For the SPARK replication, we limited our analyses to the 51 unique recurrent DELs (**Supplementary Table 13**) and 51 unique recurrent DUPs (**Supplementary Table 14**) we observed across the three defined SPARK ancestry groups.

### Carrier Prevalence

We calculated carrier prevalence as the number of individuals within a given ancestry group who carried at least one of a given class of CNV divided by the total number of individuals in that ancestry group. For the UKBB, prevalence calculations were made for the following classes: (1) carriers of at least one DEL > 50kB, (2) carriers of at least one DUP > 50kB, (3) carriers of at least one of 50 pre-defined recurrent DELs (**Supplementary Table 2**), (4) carriers of at least one of 60 pre-defined recurrent DUPs (**Supplementary Table 3**), (5) carriers of at least one DEL with a total 1/LOEUF of at least 5.7 summed across all full genes included in all DELs plus any partial genes included in any of 50 pre-defined recurrent DELs, (6) carriers of at least one DUP with a total 1/LOEUF of at least 5.7 summed across all full genes included in all DUPs plus any partial genes included in any of 60 pre-defined recurrent DUPs, (7) carriers of at least one of the 50 pre-selected recurrent DELs with a total 1/LOEUF of at least 5.7 summed across full and partial genes included in those recurrent DELs, and (8) carriers of at least one of the 60 pre-selected recurrent DUPs with a total 1/LOEUF of at least 5.7 summed across full and partial genes included in those recurrent DUPs. Prevalence calculations for the SPARK replication were limited to (1) carriers of at least one of 51 pre-defined recurrent DELs (**Supplementary Table 13**), (2) carriers of at least one of 51 pre-defined recurrent DUPs (**Supplementary Table 14**), (3) carriers of at least one of the 51 pre-selected recurrent DELs with a total 1/LOEUF of at least 5.7 summed across full and partial genes included in those recurrent DELs, and (4) carriers of at least one of the 51 pre-selected recurrent DUPs with a total 1/LOEUF of at least 5.7 summed across full and partial genes included in those recurrent DUPs. We used a threshold of total 1/LOEUF = 5.7, which corresponds to approximately two intolerant genes, as a means of identifying carriers of deleterious CNVs.

We also evaluated ancestral differences in carrier prevalence for 5 individual recurrent DELs (*NRXN1*, 2q13 *NPHP1*, *ZNF92*, *CRYL1*, and 15q11.2) and 6 individual recurrent DUPs (1q21.1 TAR, 2q13 *NPHP1*, 15q11.2, 15q13.3 BP4.5-BP5 *CHRNA7*, 16p13.11, and 22q11.2) in the UKBB. A recurrent CNV was selected for this round of analysis if it had at least 275 total observations in the combined UKBB dataset. We chose this threshold *a priori* because, under the null hypothesis that CNV carrier prevalence is not associated with ancestry (i.e., they are equally distributed across the ancestry groups), we would expect at least five carriers of each CNV in the AFR and SAS ancestry groups. For the SPARK replication, we analyzed the same eleven recurrent CNVs we selected for analysis for the UKBB along with two additional CNVs, 16p11.2 proximal DEL and 16p12.1 DEL, that each had more than 75 observations in SPARK (since we would expect at least 5 observations of these CNVs in the AFR and AMR groups under our null hypothesis) but did not meet our inclusion criteria for the UKBB.

### Sensitivity Analysis

We repeated our carrier prevalence calculations after excluding all individuals who had a least one third-degree or closer relative in the full UKBB dataset. Since these exclusions reduced our sample sizes (**Supplementary Tables 4-6**) without altering our primary findings (**Supplementary Table 4; Extended Data Figs. 2 and 3**), all subsequent UKBB analyses were run using only the full dataset.

### Environmental Phenotypes

Next, we explored whether the observed ancestry-related differences in CNV carrier prevalence could be explained by ascertainment differences. Hypothesizing that recent immigrants may be genetically fitter and, hence, less likely to possess recurrent CNVs, we compared the birth countries (Data-field 1647) of the recurrent CNV carriers within each ancestry group to those of the non-carriers (**Extended Data Fig. 4**) and quantified the difference for each ancestry group with the ratio of the odds of being a recurrent CNV carrier when born within the UK or Ireland to the odds when born elsewhere. Individuals born in England, Wales, Scotland, Northern Ireland, or the Republic of Ireland were coded as being born within the UK or Ireland. Individuals who indicated they did not know their place of birth or preferred not to answer were excluded from this analysis.

We also considered whether Townsend Deprivation Index (TDI; Data-field 189), a proxy for socioeconomic status, differed between ancestry groups or based on recurrent CNV carrier status (**Supplementary Table 7**). TDI scores were assigned to each subject based on the postal code where they resided immediately prior to enrolling in the UKBB. Note that larger, more positive TDI scores are associated with a higher degree of deprivation.

### Propensity Score Matching

Given the unbalanced sample sizes, sex ratios, age distributions, and TDI distributions across the four ancestry groups in the UKBB (**Supplementary Tables 5 and 7**), we employed propensity-score matching to control for these potentially confounding variables before running another round of analyses. After excluding individuals who were missing age and/or TDI data, we used the MatchIt^71^ R package to make two sets of 1:1 nearest neighbor matches without replacement by estimating a propensity score via logistic regression of ancestry on TDI, age, and sex (**Extended Data Fig. 5**). One WB subgroup (WB-AFR; *n* = 8,425) was matched to the remaining AFR individuals (*n* = 8,425), and a second WB subgroup (WB-SAS; *n* = 8,835) was matched to the remaining SAS individuals (*n* = 8,835); we did not include the EUR group in this round of analyses. The matching procedure yielded good balance, as evidenced by standardized mean differences and empirical CDF statistics close to zero and variance ratios close to one for both the WB-AFR matches (**Supplementary Table 8**) and WB-SAS matches (**Supplementary Table 9**).

We used odds ratios (ORs) to quantify the differences in carrier frequency between the matched datasets. Odds were calculated as the number of carriers divided by the number of non-carriers within a given group. Then, two sets of ORs were computed by dividing the AFR odds by the WB-AFR odds and the SAS odds by the WB-SAS odds. These ORs were computed for the same four classes of recurrent CNVs and 11 individual recurrent CNVs that were described above.

The SPARK replication dataset also had confounding variables that we controlled using propensity score matching. Not only do the sex ratios and ages vary substantially between ASD cases and controls and between ancestry groups, but the relative proportions of ASD cases with and without intellectual disability (ID) also differ across the ancestry groups (**Supplementary Tables 10 and 12**). We distinguished between cases and controls using the *asd* variable from the individuals_registration file and assigned ID status to the cases using the *derived_cog_impair* variable from the predicted_iq_experimental file included in the SPARK V10 release.

After excluding individuals who were missing age or ID data, we used the MatchIt^71^ R package to make two sets of 1:1 nearest neighbor matches without replacement by estimating a propensity score via logistic regression of ancestry on combined ASD/ID status (i.e., control, ASD without ID, or ASD with ID), age, and sex (**Extended Data Fig. 7**). One EUR subgroup (EUR-AFR; *n* = 3,473) was matched to the remaining AFR individuals (*n* = 3,473), and a second EUR subgroup (EUR-AMR; *n* = 7,405) was matched to the remaining AMR individuals (*n* = 7,405). The matching procedure yielded good balance, as evidenced by standardized mean differences and empirical CDF statistics close to zero and variance ratios close to one for both the EUR-AFR matches (**Supplementary Table 15**) and EUR-AMR matches (**Supplementary Table 16**).

We used ORs to quantify the differences in carrier frequency between the matched SPARK datasets. Odds were calculated as the number of carriers divided by the number of non-carriers within a given group. Then, two sets of ORs were computed by dividing the AFR odds by the EUR-AFR odds and the AMR odds by the EUR-AMR odds. These ORs were computed for the same four classes of recurrent CNVs and the 13 individual recurrent CNVs that were described above.

### Statistical Analyses

R (version 4.2.1)^72^ was used to run statistical analyses and generate graphical displays. Carrier prevalence differences among the four unmatched UKBB ancestry groups were evaluated for statistical significance using chi-square tests for independence with simulated *p*-values (2000 iterations). All tests included an additional “none of the above” group (not included in the figures) so that a given observed proportion (i.e., observed count / *n*) was equivalent to carrier prevalence. Significant differences between expected and observed carrier counts were identified using Benjamini-Hochberg FDR-corrected *p*-values assigned to the standardized residuals. The ORs for the matched UKBB comparisons (AFR vs. WB-AFR and SAS vs. WB-SAS) and matched SPARK comparisons (AFR vs. EUR-AFR and AMR vs. EUR-AMR) were evaluated using two-sided Fisher’s exact test *p*-values and plotted with their corresponding 95% confidence intervals. We conducted a Mantel-Haenszel test of homogeneity to compare the ancestry-specific birth location ORs for the UKBB, and we used two-sided Fisher’s exact tests to evaluate whether the odds of being a recurrent CNV carrier were lower for individuals of given ancestry group who were born outside the UK or Ireland (i.e., immigrants) than they were for those born within the UK or Ireland. We plotted the Mantel-Haenszel pooled OR and ancestry-specific birthplace ORs with their 95% Wald confidence intervals.

## Data Availability

Raw genotype and phenotype data are available via application to the UK Biobank (https://www.ukbiobank.ac.uk/). Approved researchers can obtain the SPARK dataset described in this study by applying at https://base.sfari.org.

## Code Availability

Custom Python code and a detailed description of our CNV calling pipeline is provided at https://martineaujeanlouis.github.io/MIND-GENESPARALLELCNV/.

## Data Availability

All data produced in the present work are contained in the manuscript.

## Acknowledgements

This work used the UK Biobank, a major biomedical database, under application number 40980 and was funded by the National Institute of Mental Health (NIMH) grant number U01-MH119690. We are grateful to all the families in SPARK, the SPARK clinical sites, and SPARK staff, and we appreciate obtaining access to phenotypic and genetic data on SFARI Base.

## Extended Data

**Extended Data Fig. 1:**
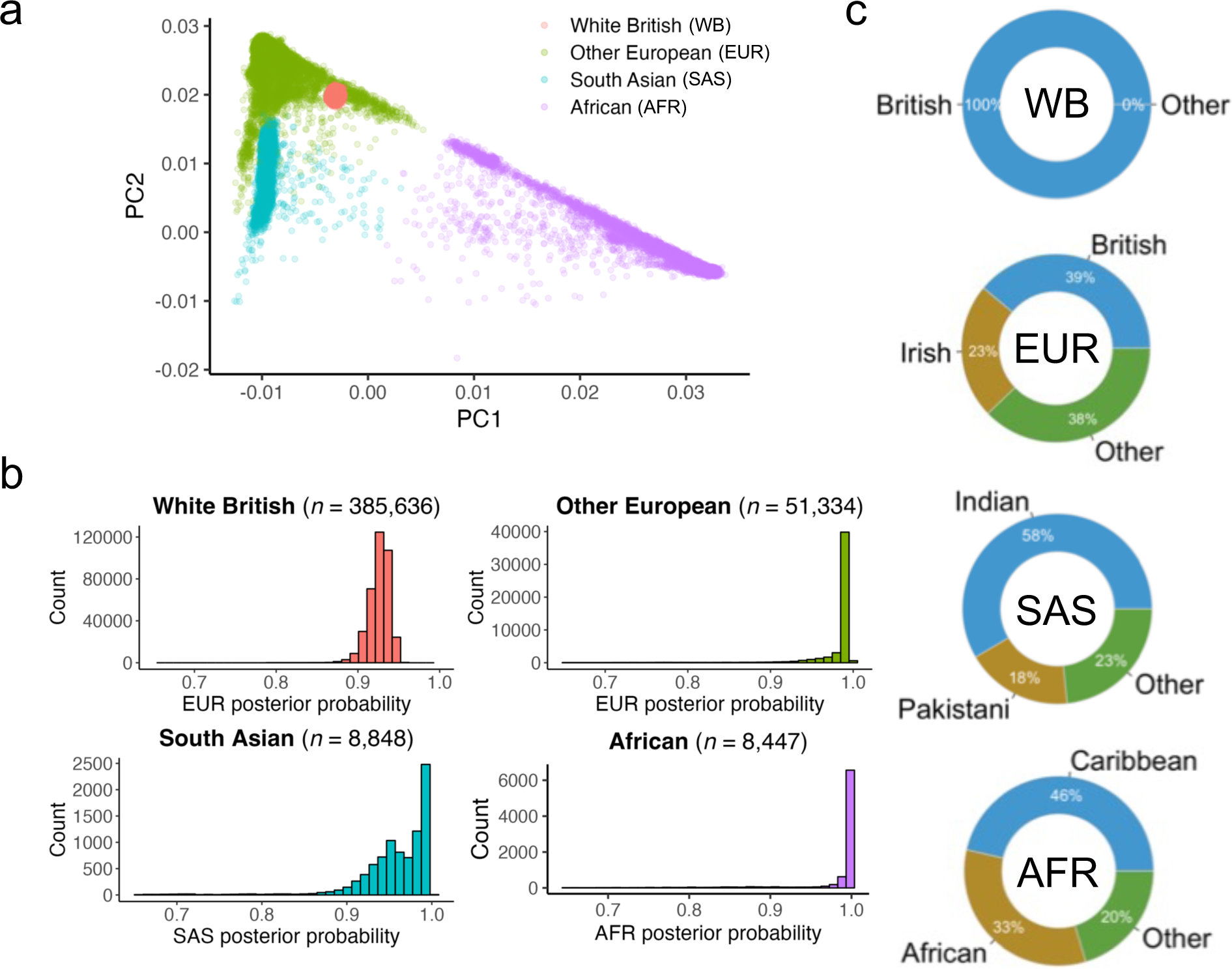
Genetic ancestry and self-identified ethnicity of UK Biobank CNV study participants. **a.** The first two ancestry principal components (PCs) are plotted with colors indicating inferred genetic ancestry. The subset of European (EUR)-ancestry individuals with self-declared white British (WB) ethnicity as defined by Data-Field 22006 is also shown. **b.** Genetic ancestry was called when the posterior probability was at least 0.65, but 93.2% of the calls were made with posterior probability ≥ 0.9. **c.** Self-declared ethnicity varied within the EUR, South Asian (SAS), and African (AFR) genetic ancestry groups; 100% of the individuals included in the WB subset of the EUR-ancestry group identified themselves as being British.

**Extended Data Fig. 2:**
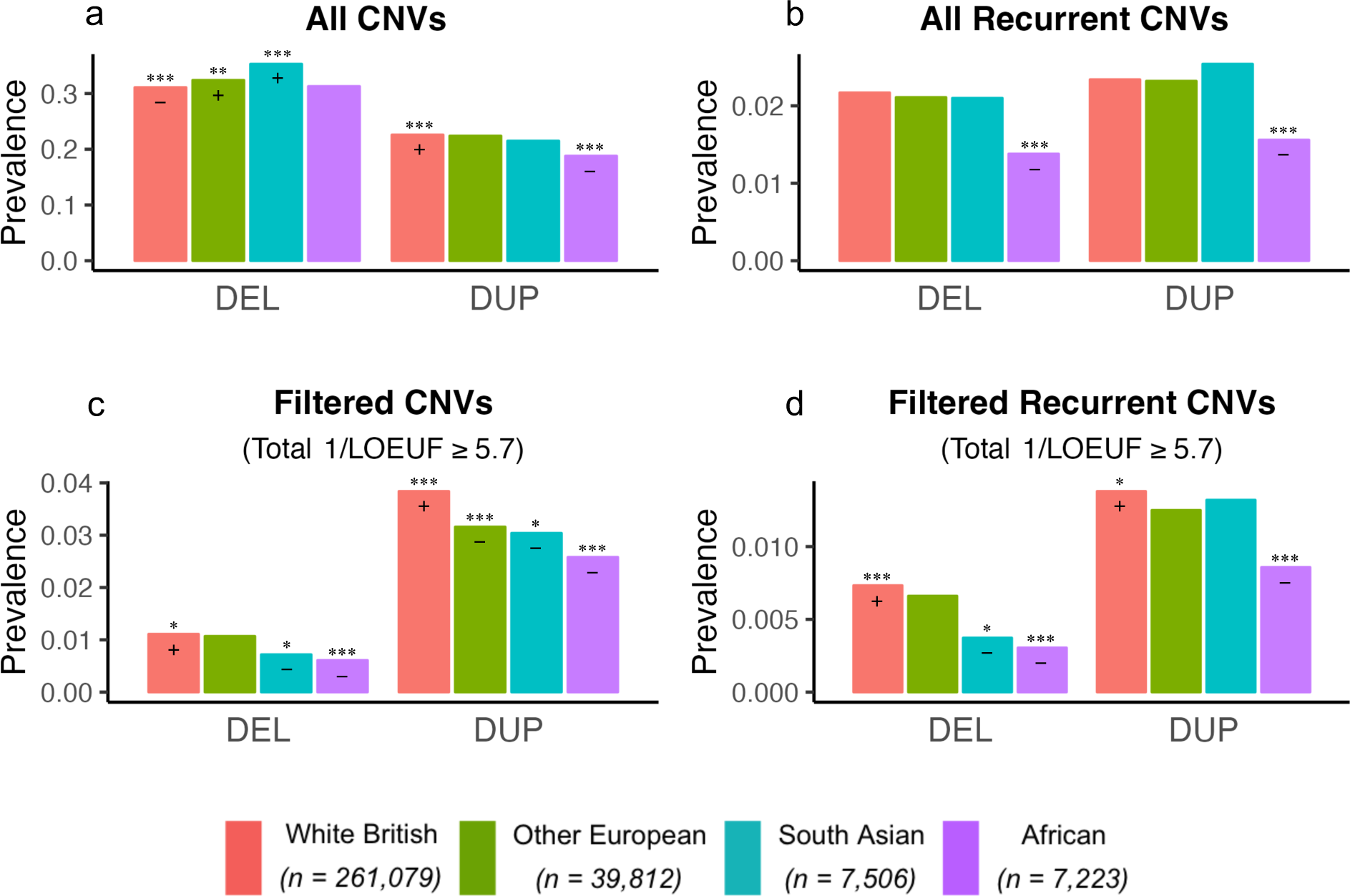
CNV carrier prevalence in the UK Biobank by ancestry after excluding all third-degree or closer relatives. These results are essentially the same to those obtained prior to excluding relatives (compare to Fig. 1). **a.** When CNVs are not filtered based on recurrence or burden, deletion (DEL) carrier prevalence is higher than expected for the South Asian ancestry (SAS) group, and duplication (DUP) carrier prevalence is lower than expected for the African ancestry (AFR) group (*χ*^2^ = 126.67). **b.** When considering only recurrent CNVs, there were fewer AFR-ancestry DEL and DUP carriers than expected (*χ*^2^ = 42.07). **c.** Filtering instead on burden (total 1/LOEUF ≥ 5.7), there were fewer DEL and DUP carriers than expected for both the AFR and SAS groups (*χ*^2^ = 108.76). **c.** Limiting to carriers of recurrent CNVs with total 1/LOEUF ≥ 5.7, DEL carrier prevalence was lower than expected for both AFR and SAS, but DUP carrier prevalence was only lower than expected for AFR (*χ*^2^ = 50.43). Plus signs indicate significantly higher than expected carrier prevalence, and minus signs indicate significantly lower than expected carrier prevalence. Ancestry-specific carrier prevalence was computed as the number of carriers of at least one DEL (or DUP) divided by the total number of individuals in that ancestry group. Simulated *p*-values (2000 replicates) were less than .0005 for all four chi-square tests of independence. 1/LOEUF, inverse loss-of-function observed/expected upper-bound fraction; *FDR-corrected *p*-value < .05; **FDR-corrected *p*-value < .005; ***FDR-corrected *p*-value < .0005. An additional category (“Neither”) that was included in each chi-square test is not shown in the bar charts.

**Extended Data Fig. 3:**
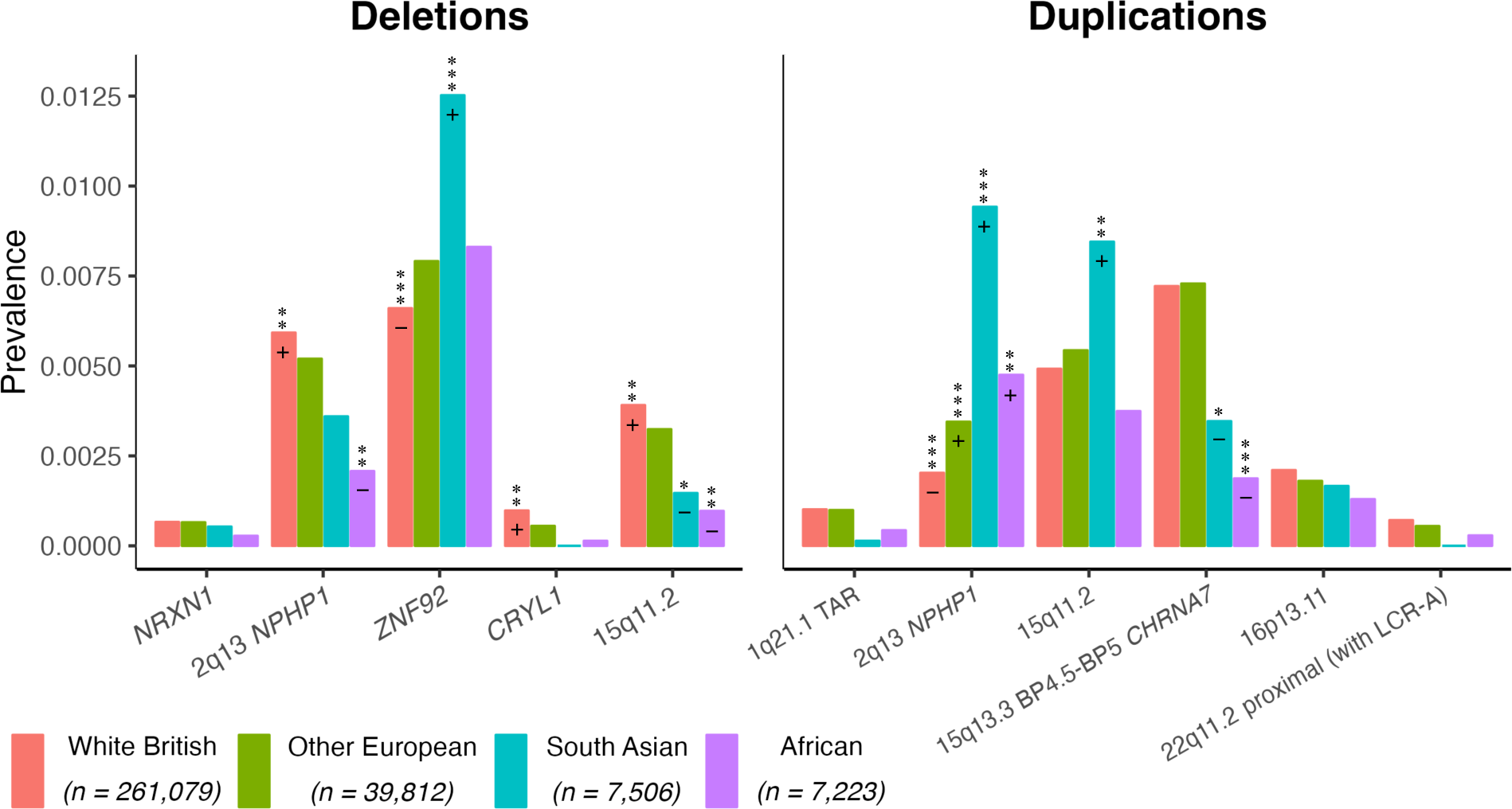
Carrier prevalence for specific recurrent CNVs by ancestry in the UK Biobank after excluding all third-degree or closer relatives. These results are essentially the same to those obtained prior to excluding relatives (compare to Fig. 2). There were significant differences between expected and observed counts for 7 out of the 11 recurrent CNVs selected for analysis, *χ*^2^ = 397.75, simulated *p*-value < .0005 (2000 replicates). Carrier prevalence was calculated as the number of carriers of a given recurrent CNV divided by the total number of individuals in that ancestry group. Plus and minus signs indicate that the standardized residuals were statistically significantly higher or lower than zero, respectively, after FDR correction. *FDR-corrected *p*-value < .05; **FDR-corrected *p*-value < .005; ***FDR-corrected *p*-value < .0005. An additional category (“none of the above”) that was included in the chi-square test is not shown in the bar chart.

**Extended Data Fig. 4:**
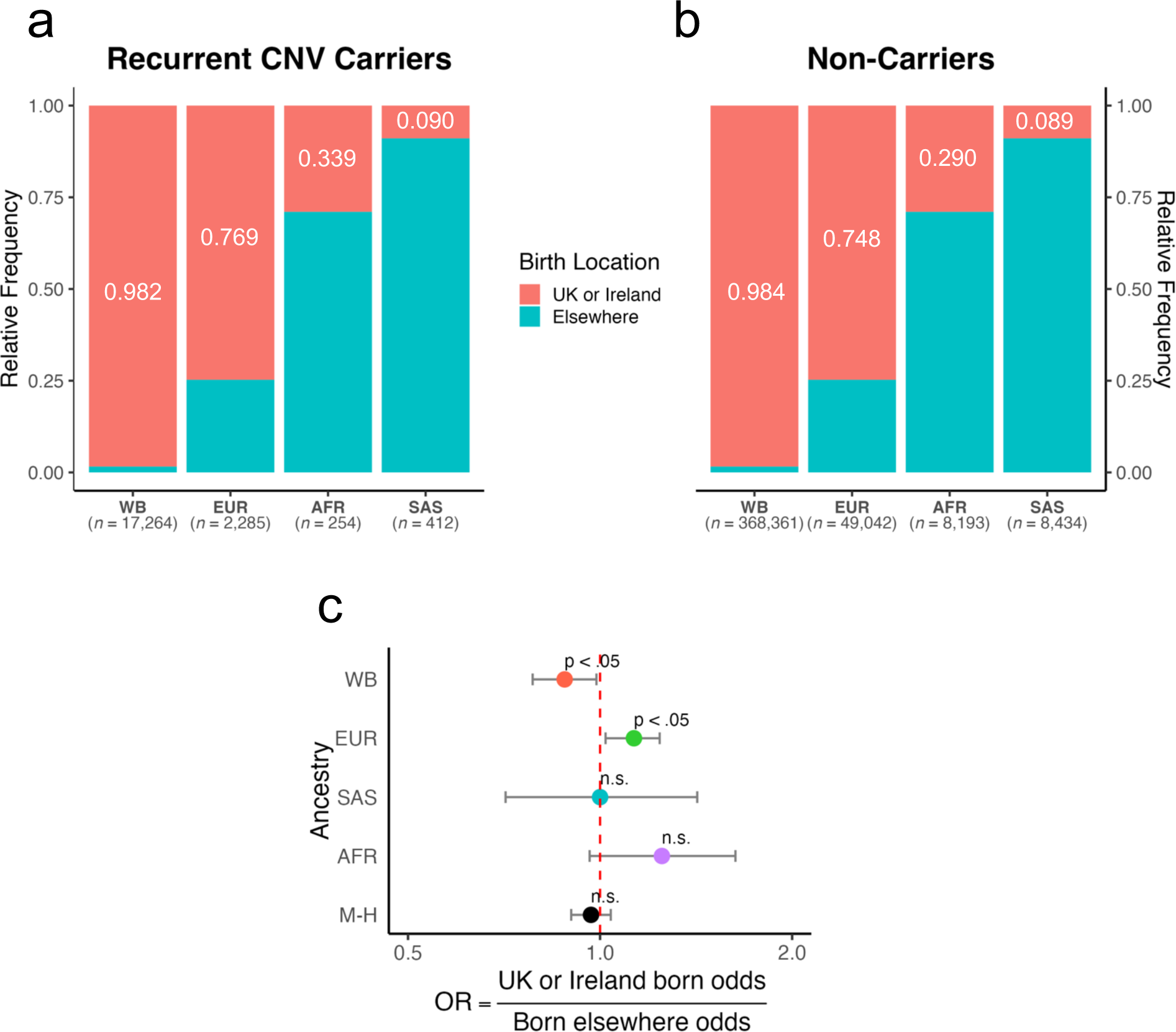
Birth locations of UK Biobank recurrent CNV carriers and non-carriers. Overall, similar proportions of recurrent CNV carriers (**a**) and non-carriers (**b**) were born in either the UK or the Republic of Ireland. (**c**) A Mantel-Haenszel test of homogeneity indicated the birth location odds ratios (ORs) were stratified by ancestry (*χ*^2^ = 12.641, *p* = .005). Ancestry-specific ORs suggest that WB recurrent CNV carriers are less likely (OR = 0.880, Fisher’s exact test *p* < .05) whereas EUR recurrent CNV carriers are more likely (OR = 1.13, Fisher’s exact test *p* < .05) to have been born in the UK or Ireland than elsewhere; the birth-location ORs for the AFR and SAS ancestry groups did not significantly differ from 1. The Mantel-Haenszel (M-H) pooled OR also did not significantly differ from 1. Note that 11 White British (WB), 7 other European (EUR), 2 South Asian (SAS), and 0 African (AFR) recurrent CNV carriers were missing birth location data; no non-carriers were missing this data. Odds were calculated as the number of recurrent CNV carriers divided by the number of non-carriers. Error bars show 95% Wald confidence limits for the ORs.

**Extended Data Fig. 5:**
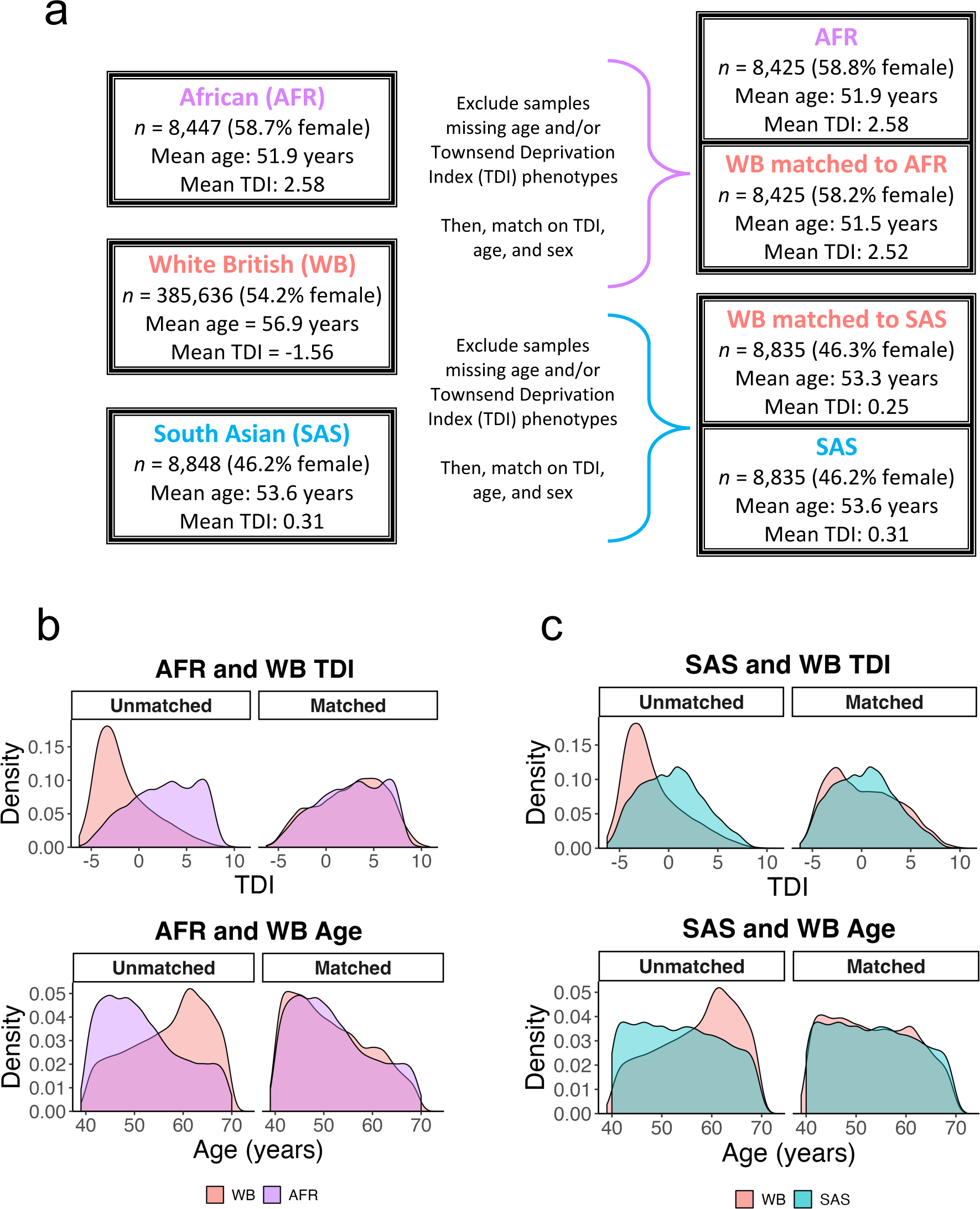
UK Biobank propensity-score matching protocol. **a.** 1:1 nearest-neighbor matching was performed after excluding subjects who were missing age and/or Townsend deprivation index (TDI) data. Two WB subsamples were matched to the filtered AFR sample (*n* = 8,425) and the filtered SAS sample (*n* = 8,848) based on TDI, age, and sex. The overlap between the density plots for TDI (top) and age (bottom) was dramatically improved between (**b**) the AFR and WB samples and (**c**) the SAS and WB samples after matching.

**Extended Data Fig. 6:**
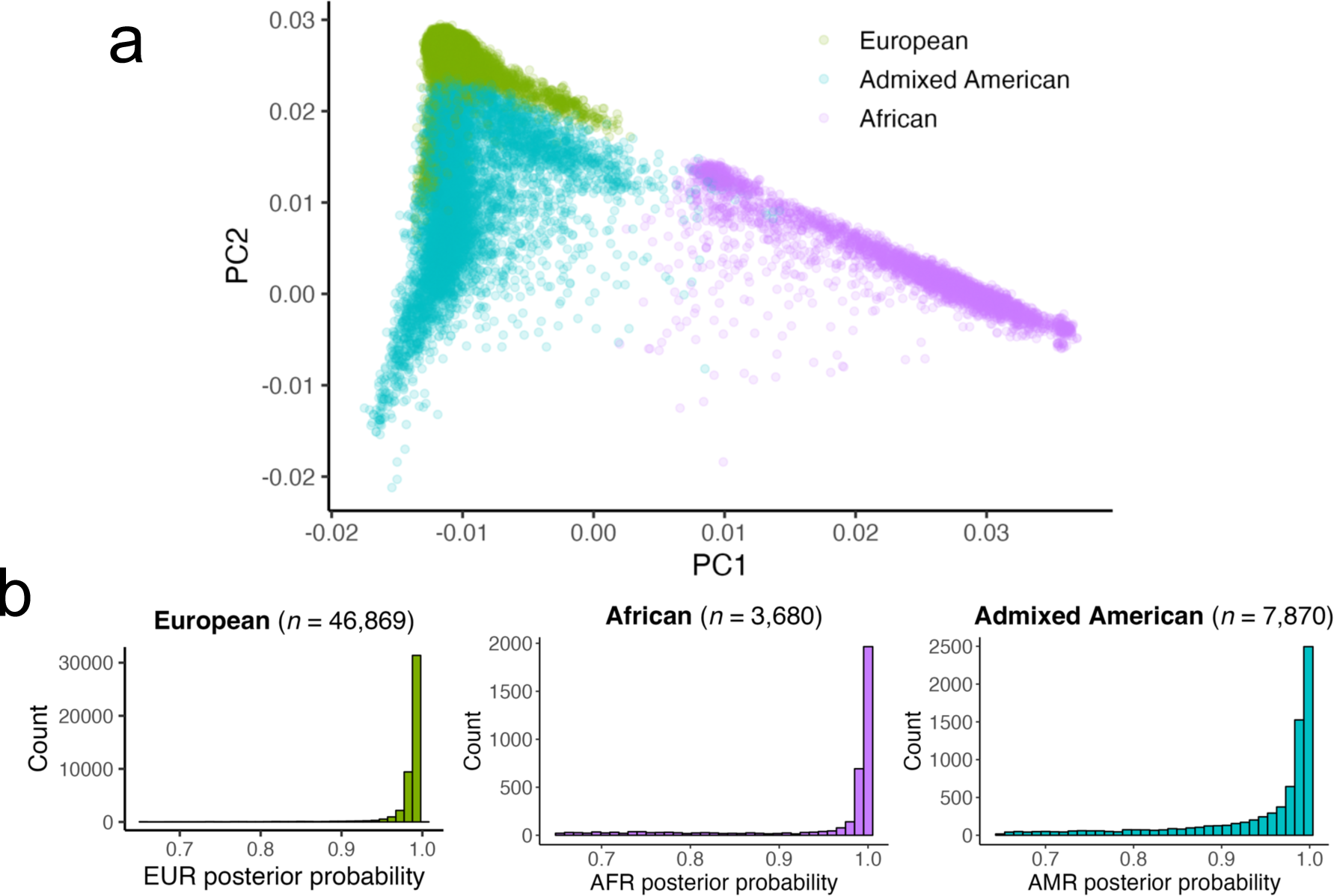
Genetic ancestry of SPARK CNV study participants. **a.** The first two ancestry principal components (PCs) are plotted with colors indicating inferred genetic ancestry. **b.** Genetic ancestry was called when the posterior probability was at least 0.65, but 96.8% of the European (EUR), 83.3% of African (AFR), and 79.4% of admixed American (AMR) ancestry calls were made with posterior probability ≥ 0.9.

**Extended Data Fig. 7:**
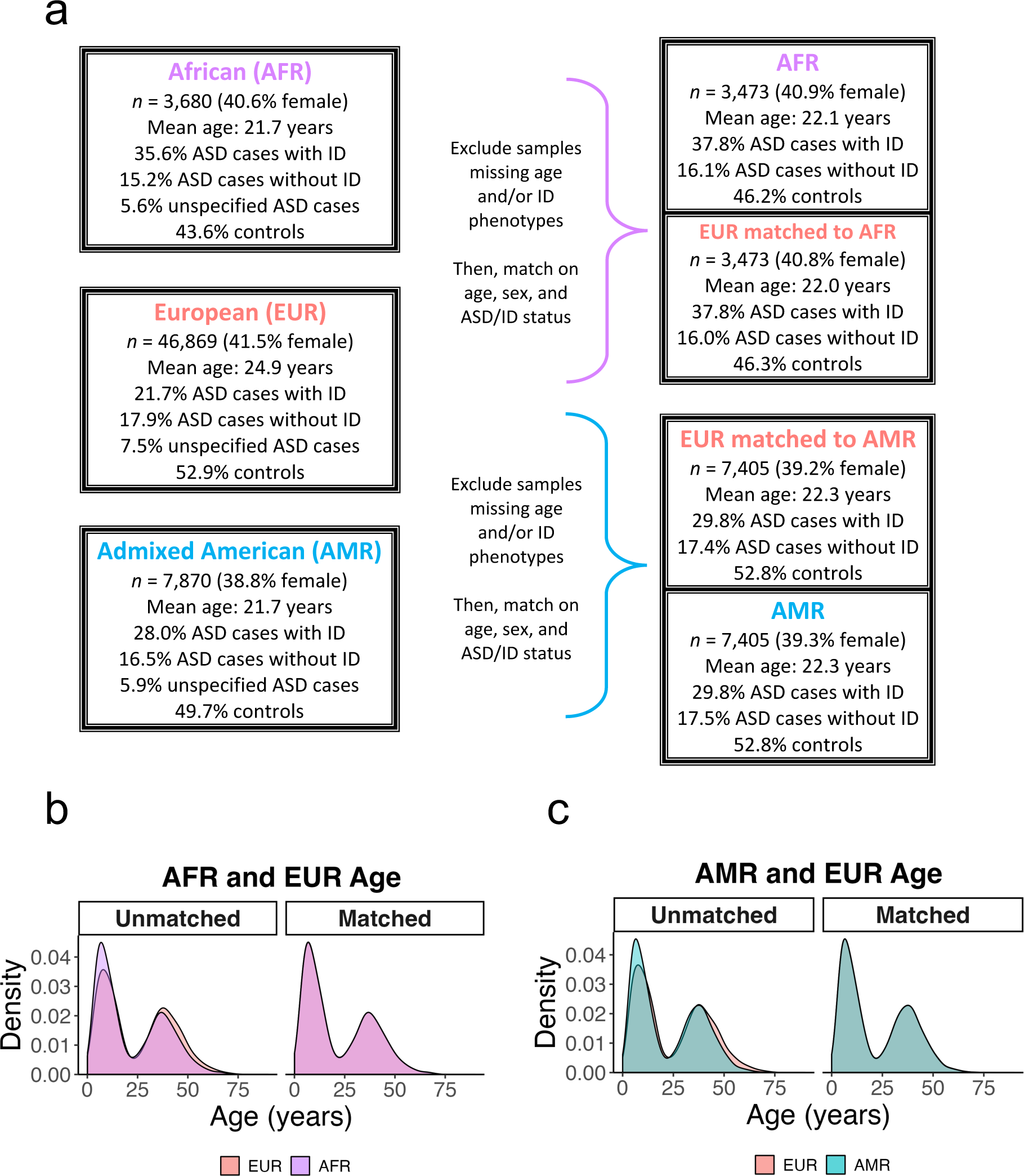
SPARK propensity-score matching protocol. **a.** 1:1 nearest-neighbor matching was performed after excluding subjects who were missing age and/or intellectual disability (ID) phenotypes. Two EUR subsamples were matched to the filtered AFR sample (*n* = 3,473) and the filtered AMR sample (*n* = 7,405) based on combined autism spectrum disorder (ASD) and ID status, age, and sex. The overlap between the density plots for age was dramatically improved between (**b**) the AFR and EUR samples and (**c**) the AMR and EUR samples after matching.

