## Supplementary Tables for "Copy number variants differ in frequency across genetic ancestry groups"

**Supplementary Table 1**: Chromosomal locations, selection criteria, affected genes, and theoretical 1/LOEUF for the autosomal recurrent and single-gene CNVs that were pre-selected for analysis

**Supplementary Table 2:** Observations of 50 unique recurrent deletions in the UK Biobank by ancestry

**Supplementary Table 3:** Observations of 60 unique recurrent duplications in the UK Biobank by ancestry

**Supplementary Table 4:** Recurrent CNV carrier status by ancestry in the UK Biobank ancestry before and after excluding all third-degree or closer relatives

**Supplementary Table 5**: Age and sex by ancestry for UK Biobank CNV study participants

**Supplementary Table 6:** UK Biobank age and sex by ancestry after excluding all third degree or closer relatives

**Supplementary Table 7:** Median Townsend deprivation index scores of UK Biobank participants by ancestry and sex

**Supplementary Table 8:** Summary of balance for propensity-score matched AFR (*n* = 8,425) and WB (*n* = 8,425) UK Biobank datasets

**Supplementary Table 9:** Summary of balance for propensity-score matched SAS (*n* = 8,835) and WB (*n* = 8,835) UK Biobank datasets

**Supplementary Table 10:** Age and sex by ancestry and ASD diagnosis for the SPARK CNV study participants

**Supplementary Table 11:** Age and sex by ancestry and ASD diagnosis for SPARK recurrent CNV carriers

**Supplementary Table 12:** Intellectual disability status of SPARK ASD cases by sex and ancestry

**Supplementary Table 13:** Observations of 51 unique recurrent deletions in SPARK by ancestry

**Supplementary Table 14:** Observations of 51 unique recurrent duplications in SPARK by ancestry

**Supplementary Table 15:** Summary of balance for propensity-score matched AFR (*n* = 3,473) and EUR (*n* = 3,473) SPARK datasets

**Supplementary Table 16:** Summary of balance for propensity-score matched AMR (*n* = 7,866) and EUR (*n* = 7,886) SPARK datasets

**Supplementary Table 1: Chromosomal locations, selection criteria, and expected 1/LOEUF for the autosomal recurrent and single-gene CNVs that were pre-selected for analysis.** These CNVs were chosen for analysis due to their previously reported associations with neurodevelopmental or neuropsychiatric phenotypes.

| Locus | Chromosome | hg19 Start Location | hg19 Stop Location | Minimal Overlap | Additional Size Criteria | Additional Gene Criteria† | Theoretical 1/LOEUF | References |
|---|---|---|---|---|---|---|---|---|
| 1p36 | chr1 | 1 | 2500000 | 40% | | *GABRD* | 75.565 | Coe et al. (2014) |
| 1q21.1 TAR | chr1 | 145394955 | 145807817 | 40% | < 1 Mb | - | 20.378 | Coe et al. (2014) |
| 1q21.1 distal + TAR | chr1 | 145394955 | 147394444 | 40% | ≥ 1 Mb | - | 30.555 | Coe et al. (2014) |
| 1q21.1 distal | chr1 | 146527987 | 147394444 | 40% | < 1 Mb | - | 9.099 | Sanders et al. (2019) |
| - | chr2 | 50145643 | 51259674 | | | *NRXN1* | 3.937 | Satterstrom et al. (2020) |
| 2q11.2 | chr2 | 96742409 | 97677516 | 40% | | *ARID5A, LMAN2L* | 43.022 | Coe et al. (2014) |
| 2q13 | chr2 | 110862716 | 110983948 | 40% | | *NPHP1* | 1.737 | Lindstrand et al. (2014) |
| 2q13 (*BUB1*) | chr2 | 111394040 | 112012649 | 40% | | - | 4.956 | Coe et al. (2014) |
| 2q21.1 | chr2 | 131481308 | 131930677 | 40% | | - | 5.807 | Coe et al. (2014) |
| 2q37 | chr2 | 239716679 | 243199373 | 40% | | *HDAC4* | 76.054 | Coe et al. (2014) |
| 3q29 proximal | chr3 | 191517306 | 193017306 | 40% | | *FGF12* | 4.737 | Coe et al. (2014) |
| 3q29 distal | chr3 | 195720167 | 197354826 | 40% | | *DLG1* | 41.321 | Coe et al. (2014); Sanders et al. (2019) |
| 4p16.3 (WH) | chr4 | 1552030 | 2091303 | 40% | | - | 21.18 | Coe et al. (2014) |
| 5q35 | chr5 | 175720924 | 177052594 | 40% | | - | 67.97 | Coe et al. (2014) |
| - | chr6 | 100836750 | 100911811 | | | *SIM1* | 3.953 | Coe et al. (2014) |
| - | chr7 | 64838768 | 64865998 | | | *ZNF92* | 0.543 | Marshall et al. (2017) |
| 7q11.23 (WBS) | chr7 | 72744915 | 74142892 | 40% | | *GTF2I, GTF2IRD1* | 50.171 | Sanders et al. (2019) |
| 7q11.23 proximal | chr7 | 73978801 | 74144177 | 40% | | - | 6.743 | Sanders et al. (2015) |
| 7q11.23 | chr7 | 74455447 | 74488775 | 40% | | *WBSCR16* | 1.044 | Sanders et al. (2015) |
| 7q11.23 distal | chr7 | 75138294 | 76064412 | 40% | | - | 16.963 | Coe et al. (2014) |
| 7p36.3 | chr7 | 158660506 | 159179546 | 40% | | *VIPR2, WDR60* | 2.456 | Vacic et al. (2011) |
| 8p23.1 | chr8 | 8098990 | 11872558 | 40% | ≥ 2 Mb | - | 25.215 | Coe et al. (2014) |
| - | chr8 | 100025494 | 100889808 | | | *VPS13B* | 1.538 | Marshall et al. (2017) |
| - | chr9 | 841690 | 969090 | | | *DMRT1* | 2.088 | Marshall et al. (2017) |
| 9q34 | chr9 | 140513444 | 140730578 | 40% | ≥ 1 Mb | *EHMT1* | 13.889 | Coe et al. (2014) |
| 10q11.21q11.23 | chr10 | 49390199 | 51058796 | 40% | | - | 21.772 | Coe et al. (2014) |
| 10q22q23 | chr10 | 82045472 | 88931651 | 40% | ≥ 1 Mb | *GRID1, NRG3* | 41.12 | Coe et al. (2014) |
| 11p11.2 | chr11 | 43940000 | 46020000 | 40% | | *EXT2* | 41.874 | Coe et al. (2014) |
| - | chr13 | 20411593 | 20437773 | | | *ZMYM5* | 0.961 | Marshall et al. (2017) |
| - | chr13 | 20977806 | 21100012 | | | *CRYL1* | 0.942 | Coe et al. (2014) |
| 13q12.12 | chr13 | 23555358 | 24884622 | 40% | | - | 8.78 | Coe et al. (2014) |
| 15q11.2 | chr15 | 22805313 | 23094530 | 40% | | - | 6.911 | Sanders et al. (2019) |
| 15q11.2q13.1 BP2-BP3 (PWS/AS) | chr15 | 22805313 | 28390339 | 40% | ≥ 4 Mb | - | 39.752 | Sanders et al. (2019) |
| 15q12 | chr15 | 26971834 | 27548820 | 40% | | *GABRA5, GABRB3, GABRG3* | 8.218 | Matsunami et al. (2013) |
| 15q13.1q13.2 BP3-BP4 | chr15 | 29161368 | 30375967 | 40% | | - | 19.614 | Stefansson et al. (2014) |
| 15q13.1q13.3 BP3-BP5 | chr15 | 29161368 | 32462776 | 40% | | - | 31.291 | Coe et al. (2014) |
| 15q13.3 BP4-BP5 | chr15 | 31080645 | 32462776 | 40% | | - | 8.692 | Sanders et al. (2019) |
| 15q13.3 BP4.5-BP5 | chr15 | 32017070 | 32453068 | 40% | | *CHRNA7* | 1.302 | Stefansson et al. (2014); Kendall et al. (2017) |
| 15q24 | chr15 | 72900171 | 78151253 | 40% | ≥ 1 Mb | - | 111.808 | Coe et al. (2014) |
| 15q25.2 | chr15 | 83219735 | 85722039 | 40% | ≥ 1 Mb | - | 45.38 | Coe et al. (2014) |
| - | chr16 | 3775056 | 3930121 | | | *CREBBP* | 15.152 | Satterstrom et al. (2020) |
| 16p13.11 | chr16 | 15511655 | 16293689 | 40% | | - | 18.381 | Sanders et al. (2019) |
| 16p12.1 | chr16 | 21950135 | 22431889 | 40% | | - | 112.305 | Coe et al. (2014) |
| 16p11.2 distal | chr16 | 28823196 | 29046783 | 40% | < 1 Mb | - | 9.033 | Sanders et al. (2019) |
| 16p11.2 distal+proximal | chr16 | 28823196 | 30200773 | 40% | ≥ 1 Mb | - | 21.389 | Coe et al. (2014) |
| 16p11.2 proximal | chr16 | 29650840 | 30200773 | 40% | < 1 Mb | - | 62.6 | Sanders et al. (2019) |
| 16p11.2p12.1 | chr16 | 21596415 | 28347808 | 40% | | - | 39.17 | Coe et al. (2014) |
| 16q23.3 | chr16 | 82660399 | 83830215 | 40% | | *CDH13* | 1.684 | Sanders et al. (2011) |
| - | chr17 | 1247834 | 1303556 | | | *YWHAE* | 4.255 | Coe et al. (2014) |
| - | chr17 | 2496923 | 2588909 | | | *PAFAH1B1* | 9.346 | Coe et al. (2014) |
| 17p12 (HNPP/CMT1A) | chr17 | 14141387 | 15426961 | 40% | | *PMP22* | 6.467 | Stefansson et al. (2014) |
| 17p11.2 (Potocki-Lupski/SMS) | chr17 | 16812771 | 20211017 | 40% | | - | 74.05 | Sanders et al. (2019) |
| 17q11.2 | chr17 | 29107491 | 30265075 | 40% | | *NF1* | 33.254 | Coe et al. (2014) |
| 17q12 | chr17 | 34815904 | 36217432 | 40% | | - | 37.084 | Sanders et al. (2019) |
| 17q21.31 | chr17 | 43705356 | 44164691 | 40% | | - | 8.334 | Coe et al. (2014) |
| 17q23.1q23.2 | chr17 | 58302389 | 60289141 | 40% | | - | 56.828 | Coe et al. (2014) |
| 22q11.2 proximal | chr22 | 19037332 | 21466726 | 40% | | | 30.023 | Sanders et al. (2019) |
| 22q11.2 distal | chr22 | 21920127 | 23653646 | 40% | | - | 74.97 | Coe et al. (2014) |
| 22q13 | chr22 | 51113070 | 51171640 | 40% | ≥ 1 Mb | *SHANK3* | 8.13 | Satterstrom et al. (2020) |

**Abbreviations:**
1/LOEUF, inverse loss-of-function observed/expected upper-bound fraction
AS, Angelman syndrome
CMT1A, Charcot-Marie-Tooth disease type 1A
HNPP, hereditary neuropathy with liabililty to pressure palsies
PWS, Prader-Willi syndrome
SMS, Smith-Magenis syndrome
TAR, thrombocytopenia-absent radius syndrome
WBS, Williams-Beuren syndrome
WH, Wolf-Hirschhorn syndrome


†Any gene(s) listed in this column needed to be disrupted for the named CNV to called. Disruption was sufficient for a single-gene DEL to be called (e.g., *NRXN1* DEL), but complete duplication was required for a single-gene DUP to be called (e.g., *NRXN1* DUP).

**Supplementary Table 2: Observations of 50 unique recurrent deletions in the UK Biobank by ancestry.**

| Recurrent Deletion | Total 1/LOEUF† | Number of Observations White British (*n* = 385,636) | Other European (*n* = 51,334) | South Asian (*n* = 8,848) | African (n = 8,447) | Total (*n* = 454,265) |
|---|---|---|---|---|---|---|
| 1q21.1 distal | 10.44 | 93 | 12 | 1 | 1 | 107 |
| 1q21.1 distal+TAR | 31.89 | 10 | 0 | 0 | 1 | 11 |
| 2q11.2 | 44.46 | 28 | 4 | 0 | 1 | 33 |
| 2q13 | 15.84 | 54 | 5 | 1 | 0 | 60 |
| **2q13 *NPHP1*** | **1.74** | **2259** | **280** | **33** | **20** | **2592** |
| 2q21.1 *BUB1* | 5.81 | 39 | 4 | 0 | 1 | 44 |
| 2q37 | 27.53 | 1 | 0 | 0 | 0 | 1 |
| 3q29 distal | 41.32 | 9 | 1 | 0 | 0 | 10 |
| 3q29 proximal | 3.84 | 3 | 0 | 0 | 0 | 3 |
| 4p16.3 | 7.18 | 5 | 1 | 0 | 0 | 6 |
| 7q11.23 distal | 20.28 | 1 | 0 | 0 | 0 | 1 |
| 7q11.23 (WBS) | 51.74 | 1 | 0 | 0 | 0 | 1 |
| 8p23.1 | 6.2 | 2 | 1 | 0 | 0 | 3 |
| 10q11.21q11.23 | 36.26 | 54 | 5 | 1 | 1 | 61 |
| 10q22q23 *NRG3 GRID1* | 44.94 | 2 | 1 | 0 | 0 | 3 |
| 13q12.12 | 8.78 | 79 | 6 | 0 | 1 | 86 |
| **15q11.2** | **6.91** | **1553** | **165** | **13** | **10** | **1741** |
| 15q11.2q13.1 BP2-BP3 (PWS/AS) | 4.98 | 1 | 0 | 0 | 0 | 1 |
| 15q13.1q13.2 BP3-BP4 | 20.13 | 9 | 1 | 0 | 0 | 10 |
| 15q13.1q13.3 BP3-BP5 | 8.69 | 47 | 7 | 1 | 2 | 57 |
| 15q13.3 BP4-BP5 *CHRNA7* | 4.31 | 0 | 1 | 0 | 0 | 1 |
| 15q13.3 BP4.5-BP5 *CHRNA7* | 1.3 | 8 | 0 | 0 | 0 | 8 |
| 15q24 | 25.47 | 0 | 1 | 0 | 0 | 1 |
| 15q25.2 | 58.1 | 0 | 1 | 0 | 0 | 1 |
| 16p11.2 distal (BP1-BP2-BP3) | 30.38 | 4 | 0 | 0 | 0 | 4 |
| 16p11.2 distal (BP2-BP3) | 21.39 | 51 | 4 | 2 | 1 | 58 |
| 16p11.2 proximal (BP4-BP5) | 40.03 | 96 | 25 | 3 | 2 | 126 |
| 16p12.1 | 9.03 | 228 | 25 | 0 | 2 | 255 |
| 16p13.11 | 24.21 | 117 | 18 | 1 | 2 | 138 |
| 16q23.3 | 2.39 | 13 | 5 | 0 | 0 | 18 |
| 17p11.2 (Potocki-Lupski/*SMS*) | 56.57 | 1 | 1 | 0 | 0 | 2 |
| 17p12 (HNPP/CMT1A) | 8.63 | 220 | 30 | 5 | 3 | 258 |
| 17q11.2 *NF1* | 35.36 | 10 | 2 | 0 | 0 | 12 |
| 17q12 | 37.08 | 6 | 2 | 0 | 0 | 8 |
| 22q11.2 distal type I | 26.67 | 4 | 0 | 0 | 0 | 4 |
| 22q11.2 distal type II | 9.57 | 40 | 6 | 0 | 0 | 46 |
| 22q11.2 distal type III | 36.58 | 1 | 0 | 0 | 0 | 1 |
| 22q11.2 proximal (with LCR-A) | 64.06 | 10 | 0 | 0 | 0 | 10 |
| 22q11.2 proximal (without LCR-A) | 20.53 | 27 | 7 | 1 | 1 | 36 |
| *CREBBP* | 19.79 | 1 | 0 | 0 | 0 | 1 |
| ***CRYL1*** | **1.52** | **361** | **36** | **0** | **1** | **398** |
| *DMRT1* | 6.07 | 13 | 2 | 0 | 1 | 16 |
| ***NRXN1*** | **3.94** | **247** | **30** | **6** | **2** | **285** |
| *PAFAH1B1* | 16.03 | 5 | 1 | 0 | 0 | 6 |
| *SIM1* | 3.95 | 5 | 0 | 0 | 0 | 5 |
| *VPS13B* | 1.54 | 33 | 3 | 2 | 0 | 38 |
| *YWHAE* | 12.98 | 4 | 0 | 0 | 0 | 4 |
| *ZMYM5* | 0.96 | 2 | 4 | 0 | 0 | 6 |
| ***ZNF92*** | **0.54** | **2641** | **408** | **106** | **77** | **3232** |
| | **TOTAL** | **8528** | **1117** | **182** | **130** | **9957** |

†Inverse loss-of-function observed/expected upper-bound fraction (1/LOEUF) was summed across all full and partial genes encompassed by a given recurrent deletion. The median 1/LOEUF score observed across all ancestries is reported.
The **boldface** rows correspond to the five deletions with more than 275 total observations; they were selected for additional analysis.

**Supplementary Table 3: Observations of 60 unique recurrent duplications in the UK Biobank by ancestry.**

| Recurrent Duplication | Total 1/LOEUF† | Number of Obervations<br>White British (*n* = 385,636) | Other European (*n* = 51,334) | South Asian (*n* = 8,848) | African (n = 8,447) | Total (*n* = 454,265) |
|---|---|---|---|---|---|---|
| 1q21.1 distal | 10.44 | 148 | 14 | 2 | 4 | 168 |
| 1q21.1 distal + TAR | 31.89 | 15 | 5 | 0 | 0 | 20 |
| **1q21.1 TAR** | **19.7** | **388** | **41** | **4** | **3** | **436** |
| 2q11.2 | 44.46 | 28 | 4 | 0 | 0 | 32 |
| 2q13 | 15.11 | 68 | 3 | 1 | 2 | 74 |
| 2q13_2q21.1 *BUB1 NPHP1* | 3.27 | 1 | 0 | 0 | 0 | 1 |
| **2q13 *NPHP1*** | **1.74** | **785** | **166** | **80** | **34** | **1065** |
| 2q21.1 *BUB1* | 5.81 | 54 | 7 | 0 | 0 | 61 |
| 2q37 | 22.56 | 1 | 0 | 0 | 0 | 1 |
| 3q29 | 23.66 | 1 | 0 | 0 | 0 | 1 |
| 3q29 distal | 43.56 | 2 | 1 | 0 | 0 | 3 |
| 3q29 proximal | 7.14 | 6 | 1 | 0 | 0 | 7 |
| 4p16.3 | 7.18 | 9 | 2 | 0 | 0 | 11 |
| 5q35 | 24.61 | 3 | 0 | 0 | 0 | 3 |
| 7q11.23 distal | 9.17 | 22 | 6 | 0 | 1 | 29 |
| 7q11.23 (WBS) | 51.74 | 13 | 2 | 2 | 0 | 17 |
| 8p23.1 | 12.75 | 6 | 0 | 1 | 0 | 7 |
| 10q11.21q11.23 | 34.27 | 38 | 4 | 0 | 1 | 43 |
| 10q22q23 *NRG3 GRID1* | 44.32 | 7 | 1 | 0 | 0 | 8 |
| 11p11.2 | 11.86 | 1 | 0 | 0 | 0 | 1 |
| 13q12.12 | 8.78 | 221 | 24 | 2 | 5 | 252 |
| **15q11.2** | **6.91** | **1849** | **278** | **69** | **30** | **2226** |
| 15q11.2q13.1 BP2-BP3 (PWS/AS) | 31.73 | 17 | 2 | 0 | 0 | 19 |
| 15q12 *GABRB3 GABRA5* | 8.22 | 7 | 1 | 0 | 0 | 8 |
| 15q13.1q13.2 BP3-BP4 | 19.08 | 20 | 3 | 1 | 0 | 24 |
| 15q13.1q13.3 BP3-BP5 | 8.69 | 214 | 29 | 4 | 1 | 248 |
| 15q13.3 BP4-BP5 *CHRNA7* | 8.69 | 56 | 8 | 0 | 1 | 65 |
| **15q13.3 BP4.5-BP5 *CHRNA7*** | **1.3** | **2660** | **351** | **31** | **18** | **3060** |
| 15q24 | 25.47 | 8 | 1 | 0 | 0 | 9 |
| 15q25.2 | 29.83 | 3 | 0 | 0 | 0 | 3 |
| 16p11.2 distal (BP1-BP2-BP3) | 31.26 | 10 | 2 | 0 | 0 | 12 |
| 16p11.2 distal (BP2-BP3) | 21.93 | 105 | 6 | 1 | 2 | 114 |
| 16p11.2 proximal (BP4-BP5) | 40.03 | 134 | 9 | 1 | 0 | 144 |
| 16p11.2p12.1 (BP1-BP2-BP3) | 147.78 | 1 | 0 | 0 | 0 | 1 |
| 16p12.1 | 9.03 | 173 | 16 | 1 | 3 | 193 |
| **16p13.11** | **23.67** | **774** | **86** | **14** | **9** | **883** |
| 16q23.3 | 2.39 | 54 | 3 | 1 | 1 | 59 |
| 17p11.2 (Potocki-Lupski/SMS) | 22.51 | 5 | 0 | 0 | 1 | 6 |
| 17p12 (*HNPP/CMT1A* ) | 8.63 | 116 | 15 | 3 | 1 | 135 |
| 17q11.2 *NF1* | 25.79 | 3 | 0 | 0 | 0 | 3 |
| 17q12 | 37.08 | 95 | 7 | 1 | 0 | 103 |
| 17q21.31 | 1.4 | 6 | 0 | 0 | 0 | 6 |
| 17q23.1q23.2 | 56.83 | 2 | 0 | 0 | 0 | 2 |
| 22q11.2 distal type I | 25.86 | 9 | 0 | 0 | 0 | 9 |
| 22q11.2 distal type II | 9.57 | 47 | 1 | 0 | 0 | 48 |
| 22q11.2 distal type III | 30.85 | 178 | 24 | 3 | 1 | 206 |
| 22q11.2 proximal distal | 178.85 | 1 | 0 | 0 | 0 | 1 |
| **22q11.2 proximal (with LCR-A)** | **76.42** | **254** | **27** | **0** | **2** | **283** |
| 22q11.2 proximal (without LCR-A) | 23.36 | 178 | 22 | 7 | 2 | 209 |
| 22q13 *SHANK3* | 64.71 | 1 | 0 | 0 | 0 | 1 |
| *CRYL1* | 3.43 | 4 | 1 | 0 | 0 | 5 |
| *CRYL1 ZMYM5* | 46.39 | 2 | 0 | 0 | 0 | 2 |
| *DMRT1* | 5.32 | 15 | 1 | 0 | 0 | 16 |
| *NRXN1* | 12.03 | 1 | 0 | 0 | 0 | 1 |
| *PAFAH1B1* | 69.86 | 1 | 0 | 0 | 0 | 1 |
| *SIM1* | 33.19 | 0 | 1 | 0 | 0 | 1 |
| *VPS13B* | 46.28 | 2 | 0 | 0 | 0 | 2 |
| *YWHAE* | 14.19 | 9 | 1 | 0 | 0 | 10 |
| *ZMYM5* | 6.48 | 66 | 12 | 0 | 0 | 78 |
| *ZNF92* | 0.54 | 202 | 19 | 7 | 6 | 234 |
| | **TOTAL** | **9099** | **1207** | **236** | **128** | **10670** |

†Inverse loss-of-function observed/expected upper-bound fraction (1/LOEUF) was summed across all full and partial genes encompassed by a given recurrent deletion. The median 1/LOEUF score observed across all ancestries is reported.

The **boldface** rows correspond to the six duplications with more than 275 total observations; they were selected for additional analysis.

**Supplementary Table 4: Recurrent CNV carrier status by ancestry for the UK Biobank before and after excluding all third-degree or closer relatives**

| Ancestry | Number of Recurrent CNVs Carried | | | |
|---|---|---|---|---|
| | 0 | 1 | 2 | 3 |
| **White British** ***n* = 385,636** (*n* = 261,079) | 0.955 (0.955) | 0.044 (0.044) | 0.0009 (0.0009) | 0.00001 (0.00002) |
| **Other European** ***n* = 51,334** (*n* = 39,812) | 0.955 (0.956) | 0.044 (0.043) | 0.0008 (0.0008) | 0 (0) |
| **South Asian** ***n* = 8,848** (*n* = 7,506) | 0.953 (0.954) | 0.046 (0.046) | 0.0007 (0.0008) | 0 (0) |
| **African** ***n* = 8,447** (*n* =7,223) | 0.970 (0.971) | 0.030 (0.028) | 0.0005 (0.0006) | 0 (0) |

The proportion of individuals within each ancestry group who carried 0, 1, 2, or 3 recurrent CNVs is shown in body of the table; no individual carried more than 3 recurrent CNVs.

The 50 recurrent DELs and 60 recurrent DUPs that contributed to these counts are defined in Supplementary Tables 2 and 3, respectively.

Results after excluding all third-degree or closer relatives are shown in parentheses.

**Supplementary Table 5: Age and sex by ancestry for UK Biobank CNV study participants**

| Group | | All Individuals | | Recurrent CNV Carriers | |
|---|---|---|---|---|---|
| | | Count (%) | Mean ± SD Age (Years) | Count (%) | Mean ± SD Age (Years) |
| **White British (WB)** | Female | 208,841 (54.2) | 56.71 ± 7.92 | 9,143 (53.0) | 56.39 ± 7.98 |
| | Male | 176,795 (45.8) | 57.14 ± 8.08 | 8,121 (47.0) | 56.88 ± 8.14 |
| | Combined | **385,636** | 56.91 ± 8.00 | **17,264** | 56.62 ± 8.06 |
| **Other European (EUR)** | Female | 28,546 (55.6) | 55.40 ± 8.09 | 1,234 (54.0) | 55.38 ± 8.25 |
| | Male | 22,788 (44.4) | 55.51 ± 8.25 | 1,051 (46.0) | 55.20 ± 8.20 |
| | Combined | **51,334** | 55.45 ± 8.16 | **2,285** | 55.30 ± 8.23 |
| **South Asian (SAS)** | Female | 4,086 (46.2) | 53.35 ± 8.12 | 210 (51.0) | 53.30 ± 8.20 |
| | Male | 4,762 (53.8) | 53.74 ± 8.73 | 202 (49.0) | 54.39 ± 8.79 |
| | Combined | **8,848** | 53.56 ± 8.46 | **412** | 53.83 ± 8.50 |
| **African (AFR)** | Female | 4,959 (58.7) | 51.98 ± 7.91 | 139 (54.7) | 51.51 ± 8.03 |
| | Male | 3,488 (41.3) | 51.75 ± 8.22 | 115 (45.2) | 51.93 ± 8.17 |
| | Combined | **8,447** | 51.89 ± 8.04 | **254** | 51.70 ± 8.08 |

**Supplementary Table 6: UK Biobank age and sex by ancestry after excluding all third-degree or closer relatives**

| Group | | All Individuals | | Recurrent CNV Carriers† | |
|---|---|---|---|---|---|
| | | Count (%) | Mean ± SD Age (Years) | Count (%) | Mean ± SD Age (Years) |
| **White British (WB)** | Female | 208,841 (54.2) | 56.71 ± 7.92 | 9,143 (53.0) | 56.39 ± 7.98 |
| | Male | 176,795 (45.8) | 57.14 ± 8.08 | 8,121 (47.0) | 56.88 ± 8.14 |
| | Combined | **385,636** | 56.91 ± 8.00 | **17,264** | 56.62 ± 8.06 |
| **Other European (EUR)** | Female | 28,546 (55.6) | 55.40 ± 8.09 | 1,234 (54.0) | 55.38 ± 8.25 |
| | Male | 22,788 (44.4) | 55.51 ± 8.25 | 1,051 (46.0) | 55.20 ± 8.20 |
| | Combined | **51,334** | 55.45 ± 8.16 | **2,285** | 55.30 ± 8.23 |
| **South Asian (SAS)** | Female | 4,086 (46.2) | 53.35 ± 8.12 | 210 (51.0) | 53.33 ± 8.17 |
| | Male | 4,762 (53.8) | 53.74 ± 8.73 | 202 (49.0) | 53.70 ± 8.83 |
| | Combined | **8,848** | 53.56 ± 8.46 | **412** | 53.83 ± 8.50 |
| **African (AFR)** | Female | 4,959 (58.7) | 51.98 ± 7.91 | 139 (54.7) | 51.51 ± 8.03 |
| | Male | 3,488 (41.3) | 51.75 ± 8.22 | 115 (45.2) | 51.93 ± 8.17 |
| | Combined | **8,447** | 51.89 ± 9.04 | **254** | 51.70 ± 8.08 |

†Recurrent CNV carriers are those individuals who carry at least one of the pre-defined CNVs listed in Supplementary Table 1.

**Supplementary Table 7: Median Townsend deprivation index scores of UK Biobank participants by ancestry and sex.** Larger, more positive values correspond to higher levels of deprivation.

| Group | | All Individuals | Recurrent CNV Carriers |
|---|---|---|---|
| **White British (WB)** | Female | -2.35 | -2.19 |
| | Male | -2.34 | -2.14 |
| | Combined | **-2.35** | **-2.17** |
| **Other European (EUR)** | Female | -1.38 | -1.46 |
| | Male | -1.23 | -1.1 |
| | Combined | **-1.3** | **-1.27** |
| **South Asian (SAS)** | Female | 0.12 | 0.21 |
| | Male | 0.45 | 0.27 |
| | Combined | **0.26** | **0.22** |
| **African (AFR)** | Female | 2.74 | 2.79 |
| | Male | 3.03 | 2.67 |
| | Combined | **2.85** | **2.74** |

**Supplementary Table 8: Summary of balance for propensity-score matched AFR (*n* = 8,425) and WB (*n* = 8,425) UK Biobank datasets**

| | AFR Mean | WB Mean | Standardized Mean Difference | Variance Ratio | eCDF Mean Difference | eCDF Maximum Difference† |
|---|---|---|---|---|---|---|
| Distance | 0.09 | 0.09 | 0.0001 | 1.0003 | 0 | 0.0007 |
| TDI | 2.59 | 2.52 | 0.0182 | 0.9515 | 0.0092 | 0.0253 |
| Age (years) | 51.89 | 51.54 | 0.0435 | 1.0304 | 0.0116 | 0.0277 |
| Female proportion | 0.59 | 0.58 | 0.0116 | NA | 0.0057 | 0.0057 |

AFR, African ancestry; WB, white British; TDI, Townsend deprivation index; eCDF, empirical cumulative density function; NA, not applicable.

†The maximum eCDF difference is equivalent to the Kolmogorov-Smirnov test statistic.

Values of standardized mean differences and eCDF statistics close to zero and values of variance ratios close to one indicate good balance.

**Supplementary Table 9: Summary of balance for propensity-score matched SAS (*n* = 8,835) and WB (*n* = 8,835) UK Biobank datasets**

| | SAS Mean | WB Mean | Standardized Mean Difference | Variance Ratio | eCDF Mean Difference | eCDF Maximum Difference† |
|---|---|---|---|---|---|---|
| Distance | 0.04 | 0.04 | 0 | 0.9999 | 0 | 0.0005 |
| TDI | 0.31 | 0.25 | 0.0211 | 0.8152 | 0.0348 | 0.0672 |
| Age (years) | 53.57 | 53.32 | 0.0294 | 1.0677 | 0.0094 | 0.0241 |
| Female proportion | 0.46 | 0.46 | -0.0016 | NA | 0.0008 | 0.0008 |

SAS, South Asian ancestry; WB, white British; TDI, Townsend deprivation index; eCDF, empirical cumulative density function; NA, not applicable.

†The maximum eCDF difference is equivalent to the Kolmogorov-Smirnov test statistic.

Values of standardized mean differences and eCDF statistics close to zero and values of variance ratios close to one indicate good balance.

**Supplementary Table 10: Age and sex by ancestry and ASD diagnosis for the SPARK CNV study participants**

| Group | | All Individuals | | ASD Cases | | ASD Controls | |
|---|---|---|---|---|---|---|---|
| | | Count (%) | Mean ± SD Age (Years) | Count (%) | Mean ± SD Age (Years) | Count (%) | Mean ± SD Age (Years) |
| **European (EUR)** | Female | 19,458 (41.5) | 30.0 ± 16.1 | 5,289 (24.0) | 15.4 ± 12.2 | 14,169 (57.2) | 35.5 ± 13.8 |
| | Male | 27,411 (58.5) | 21.3 ± 16.9 | 16,788 (76.0) | 11.8 ± 8.7 | 10,623 (42.8) | 36.2 ± 15.9 |
| | Combined | **46,869** | 24.9 ± 17.1 | **22,077** | 12.7 ± 9.8 | **24,792** | 35.8 ± 14.7 |
| **Admixed American (AMR)** | Female | 3,056 (38.8) | 27.6 ± 15.6 | 811 (20.5) | 11.2 ± 9.5 | 2,245 (57.4) | 33.5 ± 12.9 |
| | Male | 4,814 (61.2) | 18.0 ± 15.4 | 3,264 (79.5) | 9.5 ± 6.4 | 1,663 (42.6) | 34.2 ± 14.4 |
| | Combined | **7,870** | 21.7 ± 16.2 | **3,962** | 9.8 ± 7.2 | **3,908** | 33.8 ± 13.5 |
| **African (AFR)** | Female | 1,494 (40.6) | 28.4 ± 15.7 | 463 (22.3) | 12.1 ± 9.7 | 1031 (64.3) | 35.8 ± 11.9 |
| | Male | 2,186 (59.4) | 17.1 ± 15.1 | 1,614 (77.7) | 10.0 ± 6.8 | 572 (35.7) | 37.3 ± 13.8 |
| | Combined | **3,680** | 21.7 ± 16.3 | **2,077** | 10.4 ± 7.6 | **1,603** | 36.3 ± 12.6 |

ASD, autism spectrum disorder

**Supplementary Table 11: Age and sex by ancestry and ASD diagnosis for SPARK recurrent CNV carriers**

| Group | | All recurrent CNV carriers | | Recurrent CNV carriers with ASD | | Recurrent CNV carriers without ASD | |
|---|---|---|---|---|---|---|---|
| | | Count (%) | Mean ± SD Age (Years) | Count (%) | Mean ± SD Age (Years) | Count (%) | Mean ± SD Age (Years) |
| European (EUR) | Female | 1,177 (40.6) | 27.8 ± 15.8 | 410 (25.4) | 15.0 ± 11.6 | 767 (59.5) | 34.7 ± 13.4 |
| | Male | 1,723 (59.4) | 19.5 ± 15.8 | 1,202 (74.6) | 12.3 ± 8.8 | 521 (40.5) | 36.2 + 15.8 |
| | Combined | **2,900** | 22.9 ± 16.3 | **1,612** | 13.0 ± 9.6 | **1,288** | 35.3 ± 14.4 |
| Admixed American (AMR) | Female | 150 (36.1) | 25.9 ± 14.8 | 47 (19.8) | 11.4 ± 7.1 | 103 (57.5) | 32.5 ± 12.5 |
| | Male | 266 (63.9) | 16.9 ± 14.7 | 190 (80.2) | 9.8 ± 6.8 | 76 (42.5) | 34.6 ± 14.2 |
| | Combined | **416** | 20.1 ± 15.3 | **237** | 10.1 ± 6.9 | **179** | 33.4 ± 13.3 |
| African (AFR) | Female | 57 (37.3) | 27.6 ± 15.8 | 21 (22.1) | 10.8 ± 7.1 | 36 (62.1) | 37.5 ± 10.1 |
| | Male | 96 (62.7) | 14.4 ± 13.9 | 74 (77.9) | 8.7 ± 6.5 | 22 (37.9) | 33.6 ± 15.2 |
| | Combined | **153** | 19.3 ± 16.0 | **95** | 9.2 ± 6.7 | **58** | 36.0 ± 12.3 |

SPARK, Simons Powering Autism Research for Knowledge; ASD, autism spectrum disorder

**Supplementary Table 12: Intellectual disability status of SPARK ASD cases by sex and ancestry**

| Group | | All ASD Cases | ASD Cases with ID | ASD Cases without ID | ASD Cases with unknown ID status |
|---|---|---|---|---|---|
| **European (EUR)** | Female | 5,289 | 2,217 | 1,610 | 1,462 |
| | Male | 16,788 | 7,975 | 6,782 | 2,031 |
| | Combined | **22,077** | **10,192 (46.2%)** | **8,392 (38.0%)** | **3,493 (15.8%)** |
| **Admixed American (AMR)** | Female | 811 | 437 | 233 | 141 |
| | Male | 3,264 | 1,766 | 1,063 | 322 |
| | Combined | **3,962** | **2,203 (55.6%)** | **1,296 (32.7%)** | **463 (11.7%)** |
| **African (AFR)** | Female | 463 | 271 | 117 | 75 |
| | Male | 1,614 | 1,040 | 442 | 132 |
| | Combined | **2,077** | **1,311 (63.1%)** | **559 (26.9%)** | **207 (10.0%)** |

**Supplementary Table 13: Observations of 51 unique recurrent deletions in SPARK by ancestry.**

| Recurrent Deletion | Total Observations | | | |
|---|---|---|---|---|
| | European (*n* = 46,869) | Admixed American (*n* = 7,870) | African (*n* = 3,680) | Total |
| 1p36 | 2 | 0 | 0 | 2 |
| 1q21.1 distal | 31 | 0 | 1 | 32 |
| 1q21.1 distal + TAR | 2 | 0 | 1 | 3 |
| 1q21.1 TAR | 22 | 1 | 0 | 23 |
| 2q11.2 | 25 | 19 | 0 | 44 |
| 2q13 | 11 | 1 | 1 | 13 |
| **2q13 *NPHP1*** | **247** | **40** | **8** | **295** |
| 2q21.1 | 7 | 1 | 0 | 8 |
| 4p16.3 | 1 | 0 | 0 | 1 |
| 5q35 | 0 | 1 | 0 | 1 |
| 7p36.3 *VIPR2 WDR60* | 0 | 1 | 0 | 1 |
| 7q11.23 distal | 2 | 2 | 0 | 4 |
| 7q11.23 proximal | 1 | 0 | 0 | 1 |
| 7q11.23 (WBS) | 6 | 0 | 0 | 6 |
| 8p23.1 | 2 | 0 | 0 | 2 |
| 10q11.21q11.23 | 9 | 0 | 0 | 9 |
| 10q22q23 *NRG3 GRID1* | 1 | 0 | 1 | 2 |
| 13q12.12 | 13 | 0 | 0 | 13 |
| **15q11.2** | **343** | **29** | **16** | **388** |
| 15q11.2q13.1 BP2-BP3 (PWS/AS) | 1 | 0 | 0 | 1 |
| 15q13.1q13.2 BP3-BP4 | 2 | 0 | 1 | 3 |
| 15q13.1q13.3 BP3-BP5 | 55 | 4 | 2 | 61 |
| 15q13.3 BP4-BP5 | 2 | 0 | 0 | 2 |
| 15q13.3 BP4.5-BP5 | 4 | 0 | 0 | 4 |
| 15q24 | 2 | 0 | 0 | 2 |
| 15q25.2 | 1 | 0 | 0 | 1 |
| 16p11.2 distal | 21 | 4 | 0 | 25 |
| 16p11.2 distal proximal | 2 | 0 | 0 | 2 |
| **16p11.2 proximal** | **59** | **10** | **8** | **77** |
| **16p12.1** | **65** | **11** | **12** | **88** |
| 16p13.11 | 15 | 6 | 0 | 21 |
| 16q23.3 | 4 | 1 | 0 | 5 |
| 17p11.2 | 1 | 0 | 0 | 1 |
| 17p12 | 31 | 3 | 0 | 34 |
| 17q11.2 NF1 | 2 | 0 | 1 | 3 |
| 17q12 | 17 | 4 | 1 | 22 |
| 17q21.31 | 1 | 0 | 0 | 1 |
| 17q23.1q23.2 | 1 | 0 | 0 | 1 |
| 22q11.2 distal | 1 | 0 | 0 | 1 |
| 22q11.2 distal type I | 2 | 0 | 0 | 2 |
| 22q11.2 distal type II | 7 | 1 | 0 | 8 |
| 22q11.2 proximal (with LCR-A) | 24 | 3 | 0 | 27 |
| 22q11.2 proximal (without LCR-A) | 8 | 0 | 2 | 10 |
| ***CRYL1*** | **43** | **3** | **0** | **46** |
| *DMRT1* | 0 | 1 | 0 | 1 |
| ***NRXN1*** | **138** | **14** | **1** | **153** |
| *PAFAH1B1* | 1 | 0 | 0 | 1 |
| *VPS13B* | 15 | 1 | 0 | 16 |
| *YWHAE* | 0 | 1 | 0 | 1 |
| *ZMYM5* | 2 | 0 | 0 | 2 |
| ***ZNF92*** | **277** | **43** | **16** | **336** |
| TOTAL | 1529 | 205 | 72 | 1806 |

The **boldface** rows correspond to the seven deletions with more than 275 total observations in UK Biobank and/or more than 75 total observations in SPARK; they were selected for additional analysis.

**Supplementary Table 14: Observations of 51 unique recurrent duplications in SPARK by ancestry.**

| Recurrent Deletion | Number of Observations | | | |
|---|---|---|---|---|
| | European (*n* = 46,869) | Admixed American (*n* = 7,870) | African (*n* = 3,680) | Total |
| 1q21.1 distal | 57 | 7 | 5 | 69 |
| 1q21.1 distal + TAR | 9 | 2 | 0 | 11 |
| **1q21.1 TAR** | **66** | **1** | **1** | **68** |
| 2q11.2 | 0 | 1 | 0 | 1 |
| 2q13 | 3 | 0 | 0 | 3 |
| **2q13 *NPHP1*** | **72** | **12** | **17** | **101** |
| 2q21.1 | 10 | 1 | 1 | 12 |
| 2q37 | 2 | 0 | 0 | 2 |
| 4p16.3 | 2 | 0 | 0 | 2 |
| 5q35 | 1 | 0 | 0 | 1 |
| 7p36.3 *VIPR2 WDR60* | 11 | 0 | 0 | 11 |
| 7q11.23 distal | 2 | 0 | 0 | 2 |
| 7q11.23 (WBS) | 8 | 5 | 2 | 15 |
| 8p23.1 | 4 | 0 | 1 | 5 |
| 10q11.21q11.23 | 4 | 1 | 0 | 5 |
| 13q12.12 | 20 | 0 | 2 | 22 |
| 15q BP1-BP5 | 1 | 0 | 0 | 1 |
| **15q11.2** | **165** | **29** | **8** | **202** |
| 15q11.2q13.1 BP2-BP3 (PWS/AS) | 41 | 5 | 5 | 51 |
| 15q13.1q13.2 BP3-BP4 | 5 | 1 | 2 | 8 |
| 15q13.1q13.3 BP3-BP5 | 29 | 0 | 0 | 29 |
| 15q13.3 BP3-BP5 | 2 | 1 | 0 | 3 |
| 15q13.3 BP4-BP5 | 42 | 2 | 1 | 45 |
| **15q13.3 BP4.5-BP5** | **352** | **65** | **8** | **425** |
| 15q25.2 | 3 | 0 | 0 | 3 |
| 16p11.2 distal | 24 | 3 | 0 | 27 |
| 16p11.2 distal proximal | 0 | 1 | 0 | 1 |
| 16p11.2 proximal | 64 | 3 | 5 | 72 |
| 16p12.1 | 13 | 2 | 1 | 16 |
| **16p13.11** | **172** | **19** | **6** | **197** |
| 16q23.3 | 5 | 1 | 0 | 6 |
| 17p11.2 | 4 | 1 | 1 | 6 |
| 17p12 | 18 | 2 | 1 | 21 |
| 17q11.2 NF1 | 3 | 0 | 0 | 3 |
| 17q12 | 50 | 9 | 2 | 61 |
| 17q21.31 | 1 | 0 | 0 | 1 |
| 22q11.2 distal | 5 | 1 | 1 | 7 |
| 22q11.2 distal type I | 3 | 0 | 1 | 4 |
| 22q11.2 distal type II | 5 | 3 | 0 | 8 |
| 22q11.2 distal type III | 25 | 6 | 4 | 35 |
| 22q11.2 proximal | 2 | 0 | 0 | 2 |
| **22q11.2 proximal (with LCR-A)** | **52** | **5** | **1** | **58** |
| 22q11.2 proximal (without LCR-A) | 53 | 5 | 5 | 63 |
| *CREBBP* | 1 | 0 | 0 | 1 |
| *CRYL1* | 2 | 0 | 0 | 2 |
| *DMRT1* | 6 | 1 | 0 | 7 |
| *NRXN1* | 0 | 1 | 0 | 1 |
| *PAFAH1B1* | 1 | 0 | 0 | 1 |
| *YWHAE* | 6 | 0 | 0 | 6 |
| *ZMYM5* | 1 | 9 | 0 | 10 |
| *ZNF92* | 44 | 13 | 2 | 59 |
| TOTAL | 1471 | 218 | 83 | 1772 |

The **boldface** rows correspond to the six duplications with more than 275 total observations in UK Biobank and/or more than 75 total observations in SPARK; they were selected for additional analysis.

**Supplementary Table 15: Summary of balance for propensity-score matched AFR ($n$ = 3,473) and EUR ($n$ = 3,473) SPARK datasets**

| | AFR Mean | EUR Mean | Standardized Mean Difference | Variance Ratio | eCDF Mean Difference | eCDF Maximum Difference† | Standardized Pair Distance |
|---|---|---|---|---|---|---|---|
| Distance | 0.08 | 0.08 | 0 | 1 | 0 | 0.0006 | 0 |
| Age at registration (years) | 22.09 | 22.04 | 0.0032 | 1.0026 | 0.0007 | 0.0017 | 0.0032 |
| Proportion ASD cases with ID | 0.38 | 0.38 | 0 | NA | 0 | 0 | 0 |
| Proportion ASD cases without ID | 0.16 | 0.16 | 0.0031 | NA | 0.0012 | 0.0012 | 0.0031 |
| Proportion controls | 0.46 | 0.46 | -0.0023 | NA | 0.0012 | 0.0012 | 0.0023 |
| Proportion female | 0.41 | 0.41 | 0 | NA | 0.0006 | 0.0006 | 0.0012 |

AFR, African ancestry; EUR, European ancestry; eCDF, empirical cumulative density function; ASD, autism spectrum disorder; ID, intellectual disability; NA, not applicable.

†The maximum eCDF difference is equivalent to the Kolmogorov-Smirnov test statistic.

Values of standardized mean differences and eCDF statistics close to zero and values of variance ratios close to one indicate good balance.

**Supplementary Table 16: Summary of balance for propensity-score matched AMR (*n* = 7,866) and EUR (*n* = 7,886) SPARK datasets**

| | AMR Mean | EUR Mean | Standardized Mean Difference | Variance Ratio | eCDF Mean Difference | eCDF Maximum Difference† | Standardized Pair Distance |
|---|---|---|---|---|---|---|---|
| Distance | 0.15 | 0.15 | 0 | 1 | 0 | 0.0004 | 0 |
| Age at registration (years) | 21.73 | 21.72 | 0.0006 | 0.9931 | 0.0006 | 0.0019 | 0.0039 |
| ASD case proportion | 0.5 | 0.5 | -0.0008 | NA | 0.0004 | 0.0004 | 0.0064 |
| Female proportion | 0.39 | 0.39 | -0.0018 | NA | 0.0009 | 0.0009 | 0.0123 |

AMR, admixed American ancestry; EUR, European ancestry; eCDF, empirical cumulative density function; ASD, autism spectrum disorder; NA, not applicable.

†The maximum eCDF difference is equivalent to the Kolmogorov-Smirnov test statistic.

Values of standardized mean differences and eCDF statistics close to zero and values of variance ratios close to one indicate good balance.
